# Could masks curtail the post-lockdown resurgence of COVID-19 in the US?

**DOI:** 10.1101/2020.07.05.20146951

**Authors:** Calistus N. Ngonghala, Enahoro Iboi, Abba B. Gumel

**Affiliations:** Department of Mathematics, University of Florida, Gainesville, FL 32611, USA; Emerging Pathogens Institute, University of Florida, Gainesville, FL 32610, USA; School of Mathematical and Statistical Sciences, Arizona State University, Tempe, Arizona, 85287, USA; Department of Mathematics and Applied Mathematics, University of Pretoria, Pretoria 0002, South Africa

**Keywords:** Mathematical model for COVID-19, community lockdown lifting, face mask compliance, diagnostic testing and detection contact tracing, self-isolation

## Abstract

The community lockdown measures implemented in the United States from late March to late May of 2020 resulted in a significant reduction in the community transmission of the COVID-19 pandemic throughout the country. However, a number of US states are currently experiencing an alarming post-lockdown resurgence of the pandemic, triggering fears for a devastating second pandemic wave. We designed a mathematical model for addressing the key question of whether or not the universal use of face masks can halt such resurgence (and possibly avert a second wave, without having to undergo another cycle of major community lockdown) in the states of Arizona, Florida, New York and the entire US. Model calibration, using cumulative mortality data for the four jurisdictions during their respective pre-lockdown and lockdown periods, show that pre-symptomatic and asymptomatically-infectious individuals are, by far, the main drivers of the COVID-19 pandemic in each of the jurisdictions. The implication of this result is that detecting and isolating individuals with clinical symptoms of the pandemic alone (even if all of them are found) may not be sufficient to effectively curtail the pandemic. To achieve such control it is crucially-necessary that pre-symptomatic and asymptomatically-infectious individuals are rapidly detected and isolated (and their contacts rapidly traced and tested). Our study highlights the importance of early implementation of the community lockdown measures. In particular, a sizable reduction in the burden of the pandemic would have been recorded in each of the four jurisdictions if the community lockdown measures were implemented a week or two earlier. These reductions are greatly augmented if the early implementation of the lockdown measures was complemented with a public face mask use strategy. It is shown that the pandemic would have been almost completely suppressed from significantly taking off if the lockdown measures were implemented two weeks earlier, and if a sizable percentage of the residents of the four jurisdictions wore face masks during the respective lockdown periods. We simulated the pandemic in the four jurisdictions under three levels of lifting of community lockdown, namely mild, moderate and high. For the scenario where the control measures adopted are at the baseline levels during the lockdown period, our simulations show that the states of Arizona and Florida will record devastating second waves of the pandemic by the end of 2020, while the state of New York and the entire US will record milder second waves. If the level of lifting of lockdown was mild (i.e., only limited community contacts and business activities are allowed, in comparison to the lockdown period), only the state of Florida will experience a second wave. Our study further shows that the severity of the projected second waves depend on the level of lifting of the community lockdown. For instance, the projected second wave for Arizona and Florida will be more severe than their first waves. It is further shown that, for high level of lifting of community lockdown measures, the increased use of face masks after the lockdown period greatly reduces the burden of the pandemic in each jurisdiction. In particular, for this high lockdown lifting scenario, none of the four jurisdictions will experience a second wave if half of their residents wear face masks consistently after their respective lockdown period. A diagnostic testing strategy that increases the maximum detection rate of asymptomatic infected individuals (followed by contact tracing and self-isolation of the detected cases) greatly reduces the burden of the pandemic in all four jurisdictions, particularly if also combined with a universal face mask use strategy. Finally, it is shown that the universal use of face masks in public, with at least moderate level of compliance, could halt the post-lockdown resurgence of COVID-19, in addition to averting the potential for (and severity of) a second wave of the pandemic in each of the four jurisdictions.

## 1 Introduction

The novel coronavirus (COVID-19) pandemic that emerged in December 2019 is, undoubtedly, the most important public health challenge facing mankind since the 1918 pandemic of influenza. The rampaging pandemic (which spread to all countries on earth), caused by a new Severe Acute Respiratory syndrome (SARS-CoV-2), continues to cause devastating public health and socio-economic impact in many parts of the world, including the US [1, 2]. The pandemic accounted for over 10.5 million confirmed cases and 513, 000 deaths by the end of June 2020 [3, 4]. Although the pandemic emerged from China, the US and Brazil bore the brunt of its public health burden (with over 2.7 million cases and 130, 122 deaths for the US; and over 1.4 million cases and 59,656 deaths for Brazil, as of June 30, 2020). In particular, the US state of New York (alone) recorded about 417, 836 confirmed cases and 32, 129 COVID-induced mortality as of June 30, 2020. Owing to the effective and sustained control measures implemented in New York state, the state (which was once the global epicenter of the pandemic) is now recording low daily new cases and COVID-related mortality (in fact, the mortality numbers now recorded in New York state is among the lowest in the US). Although many US states have been witnessing declines in disease incidence (since the lifting of lockdown measures), a number of states (notably Arizona, California, Florida, and Texas) have recently started experiencing a dramatic increase in the number of cases and hospitalizations. In particular, Arizona and Florida recorded 4, 877 new cases (on July 1, 2020) and 15, 300 new cases (on July 12, 2020), respectively. These staggering numbers are the highest daily case counts for both states since the pandemic started (with the 15, 300 new cases in Florida being the highest number of daily cases for any US state) [5]. Furthermore, ICU admissions at the Texas Medical Center in Houston City were at 97% bed capacity as of June 24, 2020 [6]. Currently, the US states of Arizona and Florida are, respectively, the top two global epicenters of the pandemic [7]. As of July 10, 2020, the two states accounted for about 116, 892 and 244, 151 confirmed cases, and 2, 082 and 4, 102 cumulative deaths, respectively). About 72, 000 new confirmed cases was reported nationwide on the same day.

The main COVID-19 transmission pathways are person-to-person transmission through respiratory droplets, and transmission through contaminated surfaces [8]. Studies have shown that some individuals infected with the novel coronavirus can be asymptomatic or symptomatic with mild, moderate, severe, or critical symptoms [9–16]. Asymptomatic individuals exhibit no COVID-19 symptoms, although they contribute in disease transmission [9]. They include exposed individuals, who are infected but do not transmit the infection; pre-symptomatic individuals, who start shedding the virus before the onset of symptoms [17]; and infectious individuals, who do not show clinical disease symptoms after the incubation period. Those with mild clinical symptoms suffer from light fever, sneezing, cough, discomfort, etc., but no pneumonia, acute respiratory distress syndrome (ARDS). These individuals do not require Intensive Care Unit (ICU) admission [9, 10]. Some of the individuals with moderate symptoms, particularly the elderly (those aged 65+) and those with pre-existing health conditions, might experience a mild form of pneumonia that require self-isolation or hospitalization, but not ARDS or ICU admission [11–14]. Clinically severe cases of COVID-19 develop symptoms that include acute respiratory distress and failure, which might damage the lungs, as well as complications requiring hospitalization and possible ventilation [10, 15]. Critical cases require ICU admission and ARDS ventilators for survival. They are at high mortality risk and generally include the elderly (those above 65 years) and people with underlying health conditions [12]. It is known that about 81% of COVID-19 confirmed cases show mild to moderate or no symptoms, 14% show severe clinical symptoms, and approximately 5% of the cases are clinically ill [9, 15]. In addition to transmission by the clinically symptomatic, pre-symptomatic and asymptomatic individuals contribute significantly to disease transmission [18–22]. This complicates control efforts, especially when they are focused only on the severely or critically ill cases. When infectious individuals show only mild to moderate or no symptoms, it is likely that they have no urge to seek medical aid or adhere to preventive measures, thereby causing negative impact on disease mitigation or containment efforts. Hence, distinguishing between various forms of disease severity is important not only in reducing community transmission (and socio-economic burden), but also in lowering the risk of infection of front-line health care workers.

Although concerted global efforts are exerted towards developing a safe and effective vaccine against COVID-19 [23–25], no such vaccine is expected to be ready for use in humans within the next three months. Furthermore, there is currently no safe and effective antiviral against the virus. Consequently, control and mitigation efforts against COVID-19 are restricted to the implementation of non-pharmaceutical interventions (NPIs). These interventions include community lockdowns, maintaining social (or physical)-distancing in public, wearing face masks in public, quarantine of suspected cases, isolation of confirmed cases, contact-tracing and testing. The success of any of these measures relies on a number of factors. For example, the success of lockdown, social-distancing, and face mask use rely on the willingness of the members of the general population to comply with the prescribed measures and the effectiveness of the measures (e.g., face mask effectiveness to prevent the transmission or acquisition of infection) [26–29]. Diagnostic (surveillance) testing relies on the ability to test as many people as possible and the sensitivity of the test in identifying cases, especially people at the early stage of the COVID-19 infection. As of July 8, 2020, the state of Arizona has tested over 641, 070 people, with a seven-day (July 2-8, 2020) average positive test at 26.8%. Similarly, the states of Florida, New York and the entire US have (as of July 8, 2020) tested over 2.3 million, 4.4 million and 37.4 million, with a corresponding seven-day test positivity rate at 19.1%, 1.1% and 8.2%, respectively. In particular, on July 8, 2020, the state of Arizona administered 12, 273 COVID-19 tests, while the states of Florida and New York tested 51, 013 and 57, 585 residents, respectively. The number of tests conducted in the whole US on July 8, 2020 was 659, 678 [30]. While the state of New York continues to have declining test positivity rate, the states of Arizona, Florida and the entire US are currently experiencing a rise in the number of positive tests for the COVID-19 pandemic [30].

Currently, the use of face masks in public is encouraged by many state and local governments [29]. Community lockdowns and social-distancing was widely implemented in most US states (these measures entail asking people to shelter at home, closing large gathering places, such as schools, malls and non-essential businesses and maintaining 6-feet distance from other humans while in public). In particular, by April 7, 2020, mandatory lockdown measures were put in place in over 42 US states [31]. While these lockdown measures and other NPIs (such as social-distancing, the use of face masks in public, personal hygiene, testing, etc.) have been effective in curtailing the spread of COVID-19 in many US states, especially the state of New York, late implementation and early relaxation of these lockdown measures seem to be having negative effects on the effort to effectively curtail the pandemic in some US states. It should be recalled that the US White House Coronavirus Task Force announced a four-phase guidelines on April 16, 2020, for states to meet before considering relaxing the community lockdowns they implemented [32]. Specifically, Phase 1 of the guidelines requires states to consider lifting lockdown measures if they (i) achieve two weeks of continuous decline in the number of COVID-19 cases, (ii) have enough testing capacity for at-risk health care workers, and (iii) have enough hospital capacity. Relaxation of lockdown measures should rely heavily on the ability of states in particular, and the nation in general, to test and contact trace as many people as possible, encourage a sizable population to use face masks consistently in public and socially (or physically)-distance (stay 6 feet apart). Although no US state met all four phases as of May 20, 2020 (in fact, it is doubtful if any state met even the first phase [33, 34]), almost all the US states started relaxing the community lockdown measures by this time. This premature relaxation or lifting of the community lockdown measures (in an effort to re-open the economy) is clearly responsible for the post-lockdown resurgence of COVID-19 in a number of states (associated with spikes in confirmed cases and hospitalizations), including the states of Arizona, California, Florida, and Texas [33, 34]. A natural question to ask, based on the current trends, is whether or not the post-lockdown resurgence will trigger a significant second wave of the pandemic in some US states, or in the entire country (and how severe the burden of the second wave might be, in comparison to the first). It is notable that some US states have started taking important measures to help halt the post-lockdown resurgence, such as pausing the re-opening phases, re-locking down some businesses, and enacting mandatory mask usage ordinances. In fact, some counties have implemented, or are contemplating, re-imposing stay-at-home orders [35–38].

In this study, we develop and use a mathematical model to address the important question of whether or not masks alone (without another cycle of major community lockdown) could curtail the post-lockdown resurgence of COVID-19 in the US. Specifically, we use the model to assess the impact of (1) early implementation of lockdown measures combined with increased mask usage; (2) varying levels of lifting of community lockdown measures (and increases in face mask compliance); and (3) detection (including tracing, testing and self-isolation) of asymptomatic infected individuals on control and mitigating the burden of the pandemic in the chosen jurisdictions. The model will also be used to predict the likelihood (and severity) of a second wave of the pandemic in the chosen jurisdictions. The model is designed and calibrated, using cumulative mortality data from the four jurisdictions, in Section 2. Theoretical analyses of the model, with respect to the asymptotic stability of its associated continuum of disease-free equilibria, is carried out in Section 3. An expression for the final size of the pandemic is also derived analytically. Numerical simulations of the model are reported in Section 4. Discussion of the main results, together with concluding remarks, are presented in Section 5.

## 2 Materials and Methods

### 2.1 Model Formulation

The objectives of this study will be achieved *via* the design, analysis, parametrization and simulations of a Kermack-McKendrick-type SEIR (susceptible-exposed-infectious-recovered) epidemic (no human demography) model for the transmission dynamics and control of COVID-19 in a population. In addition to incorporating pertinent aspects of the epidemiology of the disease, the model to be developed will allow for the assessment of the non-pharmaceutical interventions being implemented in the US, notably social-distancing, face mask usage and contact-tracing and testing. The model to be developed is based on stratifying the total human population at time *t*, denoted by *N* (*t*), into mutually exclusive compartments based on disease status. Specifically, we split *N* (*t*) into the sub-populations of individuals who are susceptible (*S*(*t*); i.e., uninfected individuals who may contract the disease at a later time), exposed (*E*(*t*); i.e., individuals who are newly-infected but are not yet infectious), pre-symptomatic exposed (*E*_*p*_(*t*); i.e., newly-infected individuals who start shedding the virus before the end of the incubation period), asymptomatically-infectious (*I*_*a*_(*t*); i.e., individuals who do not show clinical symptoms of the disease at the end of the incubation period), infectious with mild symptoms (*I*_*m*_(*t*)), infectious with severe symptoms (*I*_*s*_(*t*)), infectious in self-isolation (*I*_*i*_(*t*)), hospitalized or isolated at a health care facility (*I*_*h*_(*t*)), individuals in intensive care units (*I*_*c*_(*t*)), recovered but not tested (*R*_*u*_(*t*)) and tested recovered (*R*_*t*_(*t*)). Further, serologic (antibody) testing of recovered humans is important for determining the level of immunity to COVID-19. Thus,

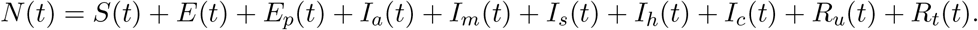

The model is given by the following deterministic system of nonlinear differential equations (where a dot represents differentiation with respect to time *t*):

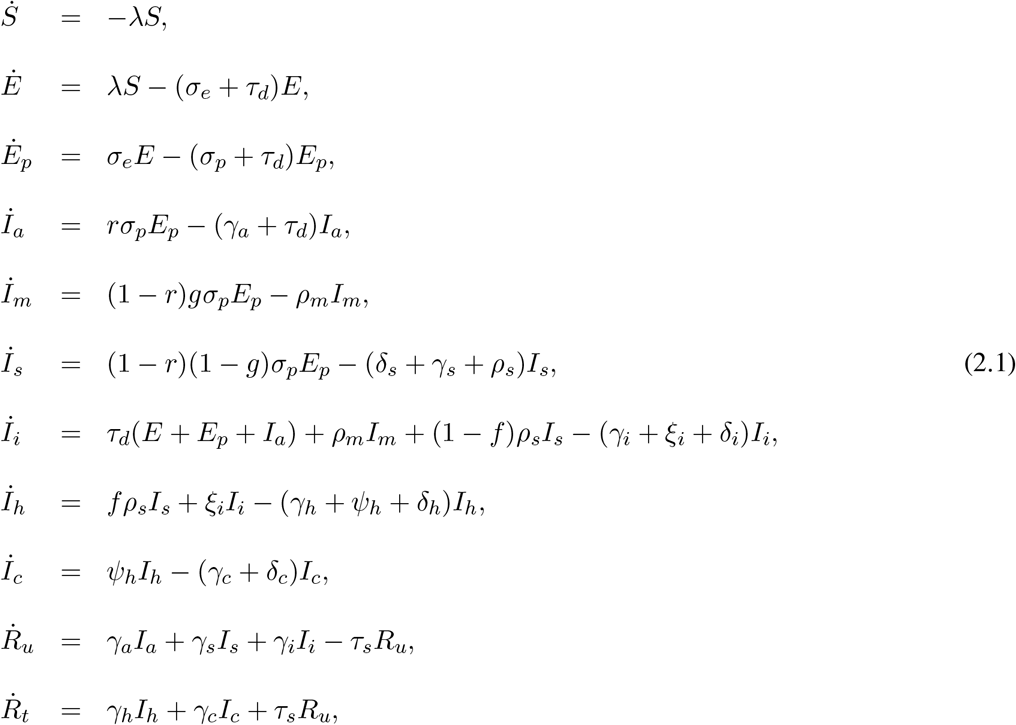

where the *force of infection λ* is defined as:

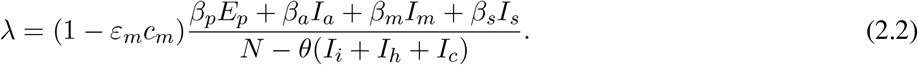

In the model (2.1), susceptible individuals acquire COVID-19 following effective contact with individuals in the pre-symptomatic (*E*_*p*_), asymptomatically-infectious (*I*_*a*_), mildly-symptomatic (*I*_*m*_) and severely-symptomatic (*I*_*s*_) classes, at a rate *λ* (defined in (2.2) below). Newly-infected individuals (in the *E* class) progress to the pre-symptomatic class at a rate *σ*_*e*_. At the end of the incubation period, a proportion, 0 < *r* ≤ 1, of humans in the *E*_*p*_ class progress to the asymptomatically-infectious class (*I*_*a*_), at a rate *rσ*_*p*_ (where 0 < *r* < 1 is the proportion of pre-symptomatic humans who do not show clinical symptoms of COVID-19 at the end of the incubation period). The remaining proportion, 1− *r*, who show clinical symptoms of COVID-19 at the end of the incubation period, are split into those who show mild symptoms (at a rate *g*(1− *r*), where 0 < *g* ≤ 1 is the proportion of the fraction), 1 − *r*, that shows mild symptoms of COVID-19) and those who show severe symptoms (at a rate (1 − *g*)(1− *r*)).

Asymptomatic individuals in the *E, E*_*p*_ and *I*_*a*_ classes are detected, *via* random diagnostic/surveillance testing, at a rate *τ*_*d*_. Similarly, serology testing is administered to untested recovered individuals (i.e., those in *R*_*u*_ class) at a rate *τ*_*s*_. Recovery in the *I*_*k*_ (*k* = *a, i, c*) class occurs at a rate *γ*_*k*_. Individuals with mild and severe symptoms are self-isolated at the rate *ρ*_*m*_ and (1− *f*)*ρ*_*s*_ (where 0 < *f*≤ 1 is the proportion of individuals with severe symptoms that are hospitalized), respectively. Self-isolated individuals are hospitalized at a rate *ξ*_*i*_. COVID-induced mortality occurs in *I*_*k*_ (*k* = *s, i, h, c*) at a rate *δ*_*k*_. Hospitalized individuals are admitted into ICU at a rate *ψ*_*h*_.

In (2.2), the term 1− *ε*_*m*_*c*_*m*_ represents a measure of the reduction in community contacts due to face mask use in the community. In particular, 0 ≤ *ε*_*m*_ ≤1 is face mask efficacy in preventing the spread (i.e., outward efficacy) or acquisition (i.e., inward efficacy) of the disease and 0 ≤*c*_*m*_ ≤ 1 is the community-wide compliance in face mask usage. The parameters *β*_*p*_, *β*_*a*_, *β*_*m*_ and *β*_*s*_ represent, respectively, the effective contact rates of infected individuals in the pre-symptomatic (*E*_*p*_), asymptomatically-infectious (*I*_*a*_), symptomatically-infectious with mild symptoms (*I*_*m*_) and symptomatically-infectious with severe symptoms (*I*_*s*_) class. The parameter 0≤ *θ*≤ 1 is a major of the effectiveness of hospitalization, self-isolation and ICU admission to prevent COVID-19 transmission by individuals in the *I*_*i*_, *I*_*h*_ and *I*_*c*_ classes.

Diagnostic and serology testing are universally considered to be highly crucial to slowing community transmission of COVID-19. In particular, diagnostic testing allows for the detection, rapid isolation and contact tracing of cases, thereby breaking the chain of community transmission that would have otherwise ensued. In the model (2.1), diagnostic testing is accounted for by way of detecting asymptomatic infected individuals, in the *E, E*_*p*_ and *I*_*a*_ classes (at the rate *τ*_*d*_). We use the following functional form for *τ*_*d*_:

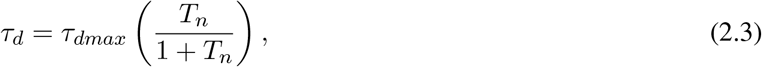

where *τ*_*dmax*_ is the maximum detection rate *via* diagnostic testing and *T*_*n*_ is the average total number of tests administered in the community *per* day. It should be noted that *T*_*n*_ can also be interpreted as a function of the number of available test kits in the community. During the 7-day period from April 30 to May 6, 2020, the average number of tests *per* day for the US was 264, 249. The maximum number of tests for the States of Arizona, Florida and New York, for the same time period, were 3, 275 (with 10.9% of these positive), 12, 223 (with 4.5% of these positive) and 22, 345 (with 13.8% of these positive), respectively [39]. These numbers are below the minimum levels recommended by the World Health Organization (WHO) (10, 743 for Arizona; 23,937 for Florida; 112,802 for New York and 917, 450 for the whole of US). The positive test ratio benchmark recommended by the WHO is 10% or less [39]. Similarly, we use the following definition for *τ*_*s*_:

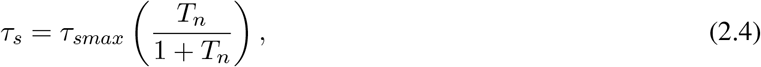

where *τ*_*smax*_ is the maximum detection rate for the serology (antibody) testing in the community.

The model (2.1) monitors human population. Hence, all its state variables are non-negative for all time *t* (further, all parameters of the model are non-negative). For housekeeping purposes, we introduce an equation for the rate of change of the population of COVID-deceased individuals (denoted by *D*(*t*)), given by:

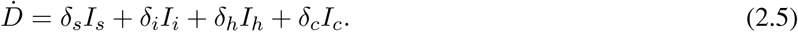

A flow diagram of the model (2.1) is depicted in Figure 1, and the state variables and parameters of the model are described in Tables A.1 and A.2 (in Appendix A), respectively.

**Figure 1:**
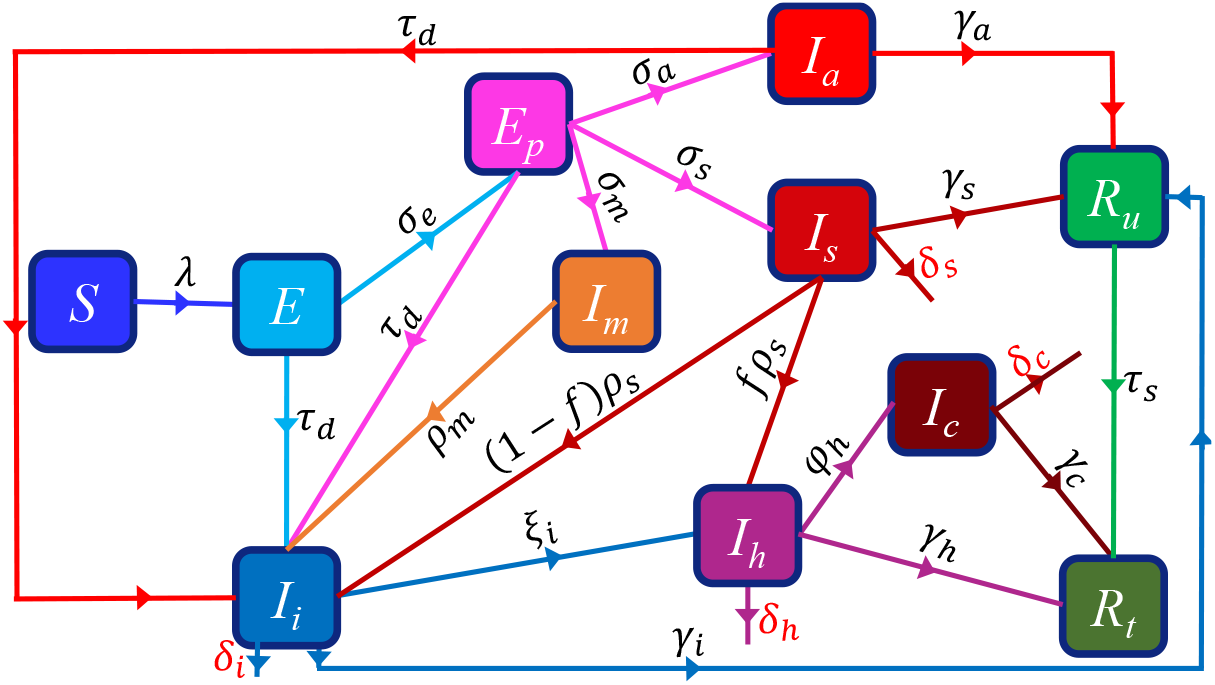
Flow diagram of the model (2.1), showing the transition of individuals between mutually-exclusive compartments based on disease status. Notation: *σ*_*a*_ = *rσ*_*p*_, *σ*_*m*_ = (1− *r*)*gσ*_*p*_, *σ*_*s*_ = (1− *r*)(1− *g*)*σ*_*p*_. The state variables and parameters are described in Table A.2.

### 2.2 Data Sources and Model Calibration

The model (2.1) has 24 parameters, and realistic values for 12 of these parameters are available in the literature (Table A.3). Estimates of the values of the remaining 12 (unknown) parameters are obtained by fitting the model to the observed cumulative deaths data for the states of Arizona, Florida, New York and the entire US. Specifically, we fit the cumulative death profile generated from the model (2.1), given by 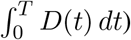 (where *T* is the prescribed future time), to the observed cumulative mortality data. Cumulative mortality data for the pre-lockdown periods for these jurisdictions (March 6 to March 31, 2020 for Arizona, March 1 to April 3, 2020 for Florida, March 1 to March 22, 2020 for New York and January 22 to April 7, 2020 for the whole of US) and the lockdown periods (March 31 to May 15, 2020 for Arizona, April 3 to May 4, 2020 for Florida, March 22 to May 28, 2020 for New York and April7 to May 28, 2020 for most of the US) was obtained from various publicly-available sources, such as the World Health Organization, the John Hopkins’ Center for Systems Science and Engineering COVID-19 Dashboard and Worldometer [5, 40–42]. The model was fitted to the data using the standard nonlinear least squares approach. This entails determining the best parameter set that minimizes the sum of the squares of the differences between Model (2.1) outputs (i.e., the model-predicted cumulative mortality) and the confirmed COVID-19 cumulative mortality data for the states of Arizona, Florida, New York, and the entire US for the pre-lockdown and lockdown periods.

The choice of fitting mortality data, as opposed the COVID-19 incidence data (which is what many COVID-19 modelers seem to prefer) is motivated by the fact that the COVID-19 mortality data is more reliable than the corresponding incidence data. The justification for this claim is that the absence of a realistic way (i.e., implementing robust rapid testing strategy across the country or jurisdiction) to quantify the size of the pool of COVID-19-infected individuals who show no symptoms of the disease makes the data for the confirmed cases to be unreliable. Lau *et al*. [43] evaluated the massive under-reporting and under-testing of COVID-19 cases in multiple global epicenters, including the US. Their data analytics study shows that, due to limited testing capacities, mortality numbers may serve as a better indicator for COVID-19 case spread in many countries (including the US). Their data indicate that countries like France, Italy, the United States, Iran, and Spain have extremely high numbers of undetected and under-reported cases. Furthermore, in a recent congressional hearing, the Director of the US Centers for Disease Control and Prevention (CDC) estimated that the current projected cumulative case data for the US may be 10 times lower than the actual case data [44].

The observed cumulative mortality data, together with the fits of the model (2.1) for the pre-lockdown and lockdown periods for the four jurisdictions, are presented in Figure 2. The estimated values of the 12 calibrated (fitted) parameters of the model (obtained from the model/data fitting) are tabulated in Tables A.4 - A.7. The baseline values of the other (12 known) parameters of the model are given in Table A.3 are drawn from the literature or estimated based on information from the literature (see [45] for details.)

**Figure 2:**
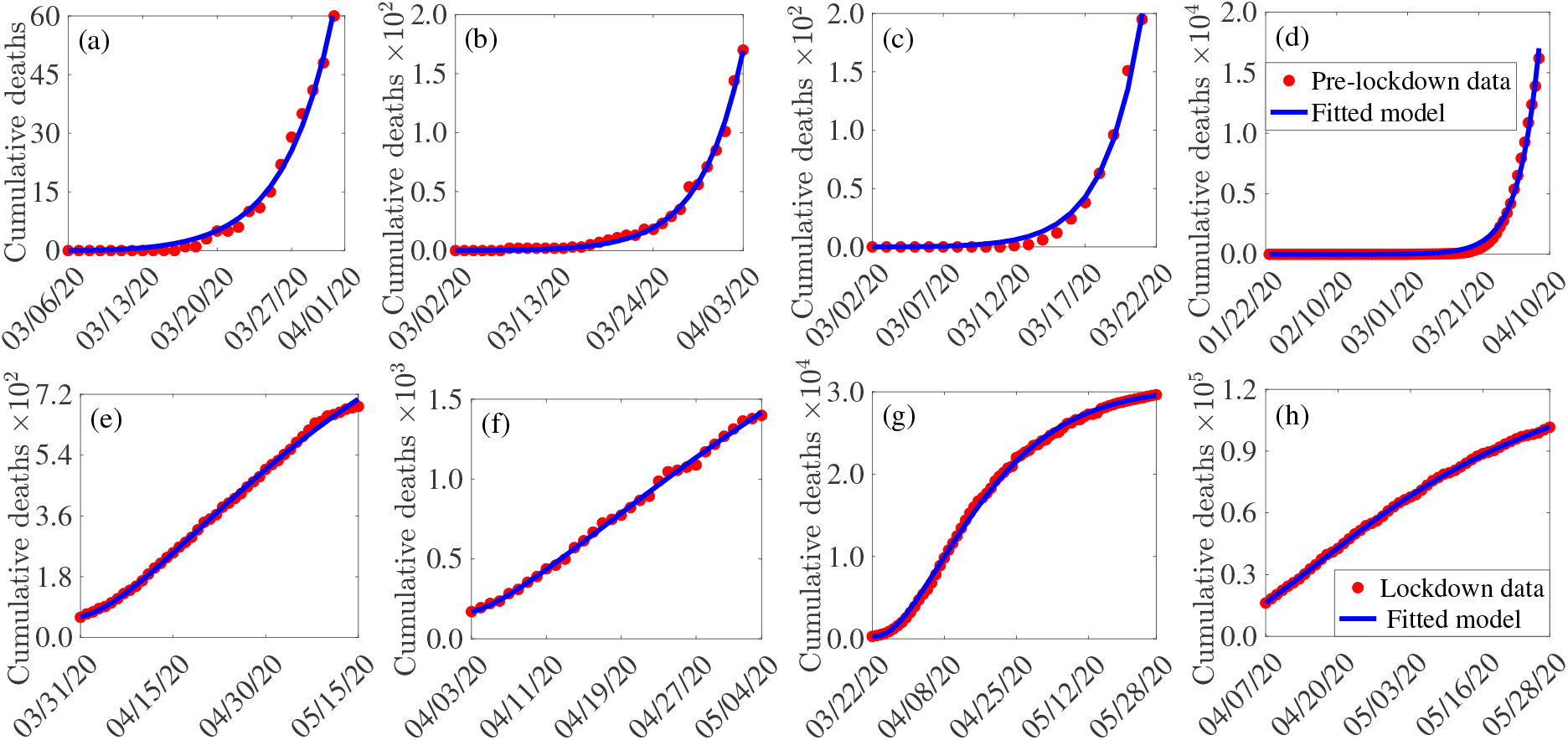
Data fitting of the model (2.1), for the pre-lockdown and lockdown periods for the states of Arizona, Florida, New York and the entire US. (a)-(d): cumulative mortality data and model fit for the pre-lockdown period for the state of Arizona, Florida, New York, and the entire US, respectively. (e)-(h): cumulative mortality data and model fit for the lockdown period for the state of Arizona, Florida, New York and the entire US, respectively.

It is evident from Tables A.4 - A.7 that the fitted values of the community contact rate parameters for the pre-symptomatic (*β*_*p*_) and asymptomatically-infectious (*β*_*a*_) individuals exceed those for the community contact rates for symptomatically-infectious individuals with mild (*β*_*m*_) and severe (*β*_*s*_) symptoms for each of the four jurisdictions (except for *β*_*m*_ during the pre-lockdown period in the state of Arizona and during the pre-lockdown and lockdown periods of the entire US; nonetheless, *β*_*p*_ + *β*_*a*_ *> β*_*m*_ + *β*_*s*_ in all scenarios, and in each of the four jurisdictions, as shown in Tables A.4 - A.7). This shows that pre-symptomatic (i.e., those in the *E*_*p*_ class) and asymptomatic (i.e., those in *I*_*a*_ class) infectious individuals are the main drivers of COVID-19 transmission in each of the four jurisdictions (this was also shown in [21, 22]. This result is further illustrated in Table A.8, showing the percentage of community transmission caused by pre-symptomatic and asymptomatically-infectious individuals (*E*_*p*_ + *I*_*a*_), in comparison to the percentage of transmission caused by individuals with mild or severe symptoms of COVID-19 (i.e., *I*_*m*_ + *I*_*s*_). Clearly, this data shows that the overwhelming majority of COVID-19 infections in each of the four jurisdiction is caused by pre-symptomatic and asymptomatically-infectious individuals. Specifically, our model calibration shows that 47-48% of COVID-19 transmission in the state of Arizona during the pre-lockdown and lockdown periods was generated by pre-symptomatic individuals. Similarly, pre-symptomatic infectious individuals account for 42-43%, 35-65%, and 25-26% of the transmissions (during the pre-lockdown and lockdown periods) in the state of Florida, state of New York, and in the entire US, respectively (see Table A.8 in Appendix A). Tindale *et al*. [21] reported that pre-symptomatic transmission played a significant role in COVID-19 dynamics in Singapore (40-50% of transmissions) and Tianjin, China (60-80%). Similar results were shown in [22] (further emphasizing the key role of pre-symptomatic and asymptomatic transmission). Our result can be intuitively explained as follows. COVID-infected individuals with no clinical (mild or severe) symptoms are unaware of their infection status, and are otherwise healthy (i.e., they are not sick). Hence, they are more likely to engage in community activities that may result in them transmitting the disease to their susceptible contacts. It is also intuitive that young people are often the ones who show no clinical symptoms of the disease after becoming infected, and such (young) people often have high community contact rates, in comparison to their older counterparts.

In conclusion, our fitting clearly shows that pre-symptomatic and asymptomatic COVID-infected individuals are the main drivers of the COVID-19 pandemic in each of the four jurisdictions. Consequently, rapidly detecting and isolating these (pre-symptomatic and asymptomatic) individuals, in addition to the rapid tracing and testing of their contacts, is crucial to the effective containment of the pandemic in each of the four jurisdictions. Furthermore, a universal face masks use strategy will certainly contribute in reducing the number of cases the pre-symptomatic and asymptomatically-infectious individuals will generate in the community.

## 3 Mathematical Analysis

### 3.1 Computation of Final Epidemic Size and Reproduction Number

The model (2.1) has a continuum (or family) of disease-free equilibria given by:

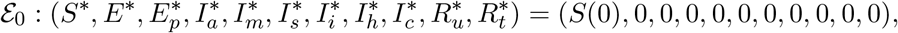

where *S*(0) is the initial number of susceptible individuals in the community. The asymptotic stability analysis of this family of equilibria will be explored below to gain qualitative insight into the behavior of initial solutions of the model, particularly with respect to the control of the burden of the pandemic. For an epidemic model (i.e., a model without demographic dynamics), such as (2.1), a useful feature to compute is the *fina lsize* of the disease [46–48]. It provides a measure of the number of individuals who remain susceptible at the end of the epidemic. The approach in [46] will be used to compute the final size relation for the model (2.1). To apply this method, it is convenient to let 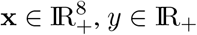, and 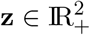 be vectors representing the compartments of infected, susceptible and recovered individuals in the model (2.1), respectively. That is, **x** = (*E, E*_*p*_, *I*_*a*_, *I*_*m*_, *I*_*s*_, *I*_*i*_, *I*_*h*_, *I*_*c*_)^*T*^, *y* = *S* and **z** = (*R*_*u*_, *R*_*t*_)^*T*^. Furthermore, suppose, Π is an 8 × 1 matrix in which the (*i, j*) entry is the fraction of the *j*^th^ susceptible compartment that goes into the *i*^th^ infected class upon becoming infected, and **b** is an − dimensional row vector of relative horizontal transmissions. It then follows from Arino *et al*. [46] that the model (2.1) reduces to the following three-dimensional system of differential equations:

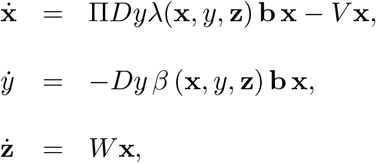

where *λ* is the *force of infection* of the model (2.1) defined in (2.2), *W* is a 2× 8 matrix in which the (*i, j*) entry is the rate at which individuals of the *j*^th^ infected compartment transition into the recovered (*i*^th^ *z*) compartment upon recovery, and *V* is the standard M-matrix of the linear transition terms between the infected compartments of the model (2.1) [46] (see also the application of next generation operator method to analyse the asymptotic stability of disease-free equilibria of disease transmission models [49, 50]). It can be seen that, in the context of model (2.1),

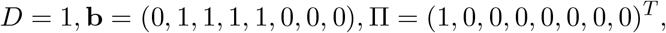

and,

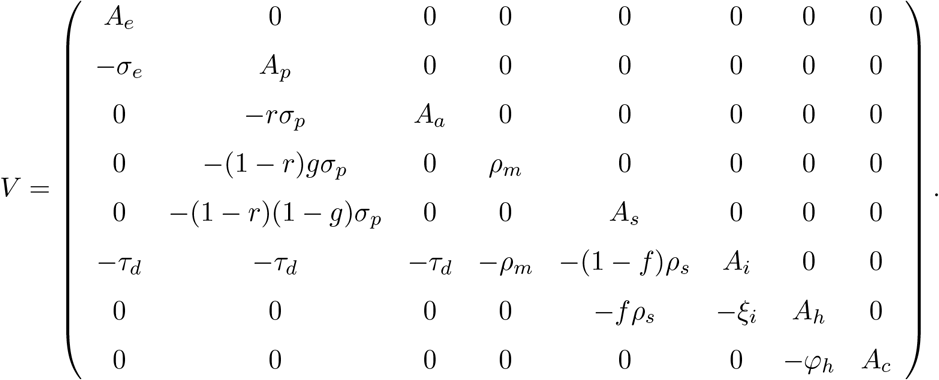

Let *y*(0) = *S*(0), **z**(**0**) = (*R*_*u*_(0), *R*_*t*_(0))^*T*^. It then follows from Theorem 2.1 in [46] that the *reproduction number* of the model (2.1), denoted by *ℛ*, is given by:

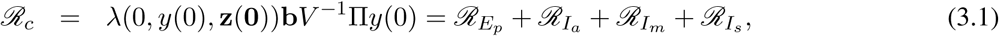

where,

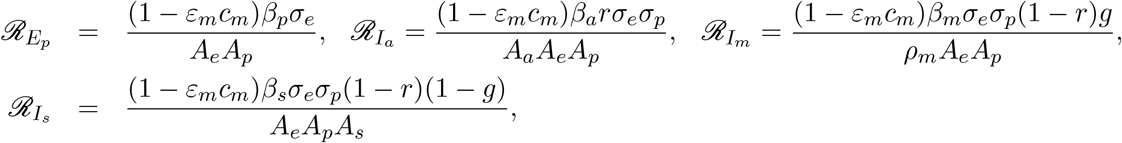

with,

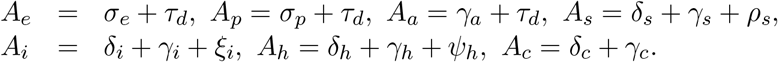

The reproduction number *ℛ*_*c*_ is an epidemiological quantity that measures the average number of new COVID-19 cases generated by a typical infected individual introduced into a community where some anti-COVID-19 interventions (such as social-distancing, face mask usage and self-isolation) are implemented. The result below follows from Theorem 2.1 of [46].

#### Theorem 3.1.

*The continuum of disease-free equilibria (ε*_0_*) of the model* (2.1) *is locally-asymptotically stable whenever ℛ*_*c*_ < 1.

The epidemiological implication of Theorem 3.1 is that community transmission of COVID-19 can be effectively suppressed in the community if the control and mitigation strategies implemented can bring the reproduction number (*ℛ*_*c*_) to a value less than unity.

#### Remark 3.1.

*For epidemic models (i*.*e*., *models with no birth and death demographic processes), such as* (2.1), *having the associated reproduction (ℛ*_*c*_*) reduced to a value less than unity is sufficient, but not necessary, for eliminating the disease. That is, even if ℛ*_*c*_ *exceeds unity, the disease eventually dies out over time. There are many reasons to explain this. First, for ℛ*_*c*_ *>* 1, *a sizable number of the population will get infected particularly if no control or mitigation interventions are implemented (or are delayed in being implemented), some of whom would survive the infection (i*.*e*., *they would recover). Since the model assumes that recovery confers perfect and permanent natural immunity against future infections, this process leads to, or contributes to, building community-wide natural herd immunity (so that many of the remaining susceptible members of the community can be protected). This, together with the implementation of control and mitigation measures, contribute in significantly suppressing community transmission, and the disease eventually dies out (even if ℛ*_*c*_ *>* 1).

Finally, it follows from Theorem 5.1 of [46] that the final epidemic size relation for the model (2.1) is given by

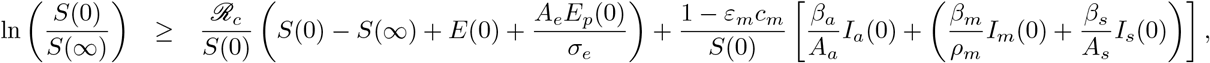

Where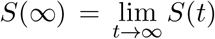. This final size relation can be solved recursively for *S*(*∞*) using, for instance, the Newton-Raphson’s method, to obtain the number of susceptible humans who remain uninfected after the epidemic.

### 3.2 Sensitivity of Reproduction Number With Respect to Case Detection and Face Mask Usage

In this section, the effect of case detection and self-isolation of asymptomatic individuals in the *E, E*_*p*_ and *I*_*a*_ classes (using diagnostic testing) and face mask usage on the reproduction number (*ℛ*_*c*_) of the model (2.1) will be assessed, for the states of Arizona, Florida, New York and the entire US. We first simulate the effect of maximum case detection (*τ*_*dmax*_) for two scenarios, where face masks of low quality (such as home-made cloth masks with estimated efficacy of 25%) or of moderate quality (such as medical/surgical masks, with estimated efficacy of 50%) are used in the community. A contour plot (heat map) of *ℛ*_*c*_, as a function of percentage increase in the maximum case detection rate (i.e., increase in the baseline value of *τ*_*dmax*_) and face mask compliance (*c*_*m*_), is depicted in Figure 3. This figure shows that the reproduction number (*ℛ*_*c*_) decreases with increasing values of the percentage increase in the maximum case detection rate and face mask compliance, for the states of Arizona (Figure 3 (a) and (e)), Florida (Figure 3 (b) and (f)), New York (Figure 3 (c) and (g)) and the entire US (Figure 3 (d) and (h)), regardless of the type (or efficacy) of the face mask used in the community. In particular, it is evident from the contour plots that the reproduction number can be brought to a value less than unity (thereby resulting in effectively curtailing community transmission of COVID-19) in Arizona, Florida and New York if the baseline maximum case detection rate is increased by at least 10%, the face mask efficacy is 25%, and face mask compliance exceeds 73%, 95%, and 35%, respectively. If the face mask efficacy is increased to 50%, then for the same maximum detection rate of 10%, face mask use compliance 37%, 48%, and 18% is required to reduce the reproduction number below one. Greater reductions in *ℛ*_*c*_ are recorded if the moderately-effective face mask (Figure 3 (e)-(h)) are used in the community, in comparison to when low effective face masks are used (Figure 3 (a)-(d)). These plots clearly show that widespread (random) testing (which then implies rapid detection, tracing and isolation of confirmed cases), combined with face mask usage, significantly contributes in reducing community transmission (by breaking the chain of transmission) regardless of the efficacy of face masks.

**Figure 3:**
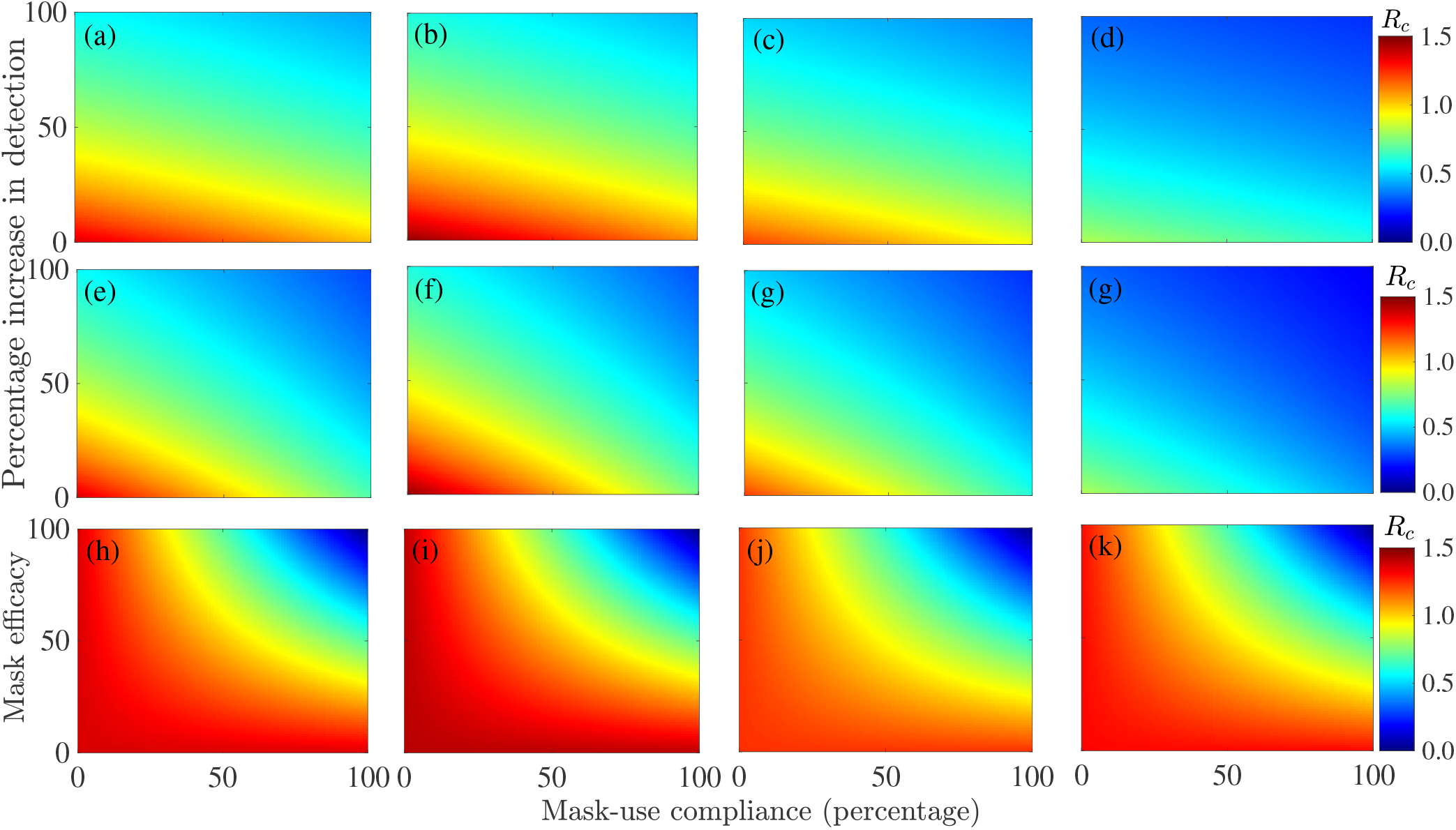
Sensitivity of the control reproduction number (*ℛ*_*c*_) to important control parameters. Heat maps of the control reproduction number, as a function of cloth face mask use compliance and percentage increases in the maximum detection rate of asymptomatic infected individuals (*τ*_*dmax*_). (a)-(d): impact of cloth masks (with efficacy *ε*_*m*_ = 0.25) on disease dynamics for the states of Arizona, Florida, New York and all of US, respectively. (e)-(h): impact of surgical/medical masks (with efficacy *ε*_*m*_ = 0.50) on disease dynamics for the state of Arizona, Florida, New York and all of US, respectively. (h)-(k): impact of face masks efficacy (*ε*_*m*_) and compliance (*c*_*m*_) on disease dynamics for the state of Arizona, Florida, New York and all of US, respectively. Parameter values used in the simulations are as given in Tables A.3-A.7 in Appendix A.

The sensitivity of the reproduction number with respect to the singular effect of face masks is monitored by generating contour plots of *ℛ*_*c*_, as a function of face mask efficacy (*ε*_*m*_) and compliance (*c*_*m*_). The results obtained, depicted in Figure 3 (i)-(l), for Arizona, Florida, New York and the entire US, show decreasing values of the reproduction number with increasing values of face mask efficacy and compliance, as expected. It is shown that, with the assumed baseline face mask efficacy of 50% (i.e., *ε*_*m*_ = 0.5), a minimum compliance of 53% will be needed to effectively curtail community transmission in the state of Arizona (Figure 3 (i)). The corresponding minimum compliance percentages needed to achieve same control in Florida, New York and the whole of US are, 59%, 38% and 46%, respectively. The contours in Figure 3 (i)-(l) also show that using low effective face masks (e.g., the cloth mask with estimated efficacy of 25%) with high compliance can lead to significant reduction in disease burden. However, using such (cloth) masks will fail to reduce the reproduction number to a value below unity, even if 100% compliance is achieved, in the states of Arizona and Florida. On the other hand, such masks can bring the reproduction number to a value less than unity for the state of New York and the entire nation if the compliance in its usage is at least 76% and 92%, respectively.

## 4 Numerical Simulations

In this section, the model (2.1) will be simulated, using the baseline parameter values in Tables A.3-A.7 in Appendix A (unless otherwise stated), to assess the community-wide impact of early implementation of strict lockdown measures (aimed at minimizing community transmission of COVID-19) in the states of Arizona, Florida, New York and the entire US. The impact of lifting of community lockdown, as well as the effect of the use of non-pharmaceutical interventions after the lifting of the lockdown, will also be assessed. Parameter values for the pre-lockdown (sub-tables (a) of Tables A.3-A.7 in Appendix A) and the lockdown period (sub-tables (b) of Tables A.3-A.7 in Appendix A) are used to simulate the COVID-19 dynamics in the four jurisdictions during the pre-lockdown and the lockdown periods, respectively. It should be mentioned that, in all of the simulations to be carried out, the control-related parameters (e.g., the face masks compliance and efficacy, *c*_*m*_ and *ε*_*m*_, and the diagnostic and serology detection rate parameters, *τ*_*d*_, *τ*_*s*_ and *τ*_*dmax*_) are kept at their baseline values, for both the pre-lockdown and lockdown periods in Tables A.3-A.7 (unless otherwise stated).

### 4.1 Impact of Early Implementation of Lockdown Measures and Mask use Compliance

It should be recalled that lockdown measures were implemented in the states of Arizona, Florida and New York on March 31, 2020, April 3, 2020 and March 22, 2020, respectively, while the entire US was on lockdown by April 7, 2020. Furthermore, partial lifting of the lockdown measures were announced in the three states by May 15, 2020, May 4, 2020 and May 28, 2020, respectively. Although some US states delayed announcing some form of lifting of lockdown measures until June 2020, the overwhelming majority of the US states implemented some form of lifting by May 28, 2020. Consequently, we assume that the entire US started partial lifting of lockdown on May 28, 2020, as was the case for the state of New York. We now run simulations of the model (2.1) for the three scenarios: (a) the precise period when community lockdown was actually implemented, (b) lockdown measures were implemented a week earlier than the actual time it was implemented, and (c) lockdown measures were implemented two weeks earlier than the actual time they were implemented, for each of the three states and the entire US.

The simulation results obtained for scenario in which lockdown was implemented one week earlier are depicted in Figure 4, showing daily and cumulative mortality for the three states and the US. This figure shows a marked decrease in the peak daily deaths in each of the four jurisdictions (Figures 4 (a)-(d), magenta curves). In other words, the pandemic curves in the three states and in the entire US would have been flattened earlier (characterized by very low numbers at the pandemic peak). The decrease in daily mortality is even more dramatic if lockdown was implemented two weeks earlier (Figures 4 (a)-(d), green curves). Furthermore, our simulations show that if the lockdown measures were implemented a week earlier in Arizona, up to 75% of the cumulative number of COVID-19 deaths in the state could have been averted, in comparison to the actual cumulative mortality recorded by the day the lockdown was partially lifted (Figure 4 (e), magenta curve). Similarly, 76% of the cumulative deaths in the state of Florida (Figure 4 (f), magenta curve) and 99% of the cumulative mortality in the state of New York (Figure 4 (f), magenta curve) would have been prevented. At least 77% of the cumulative deaths recorded in the entire US would have been averted if the lockdown measures were implemented a week earlier, in comparison to the actual cumulative mortality recorded by the day the lockdown measures were partially lifted in the US (Figure 4 (h), magenta curve). More dramatic reductions in cumulative mortality would have been recorded if the lockdown measures were implemented two weeks earlier (Figure 4 (e)-(h), green curves).

**Figure 4:**
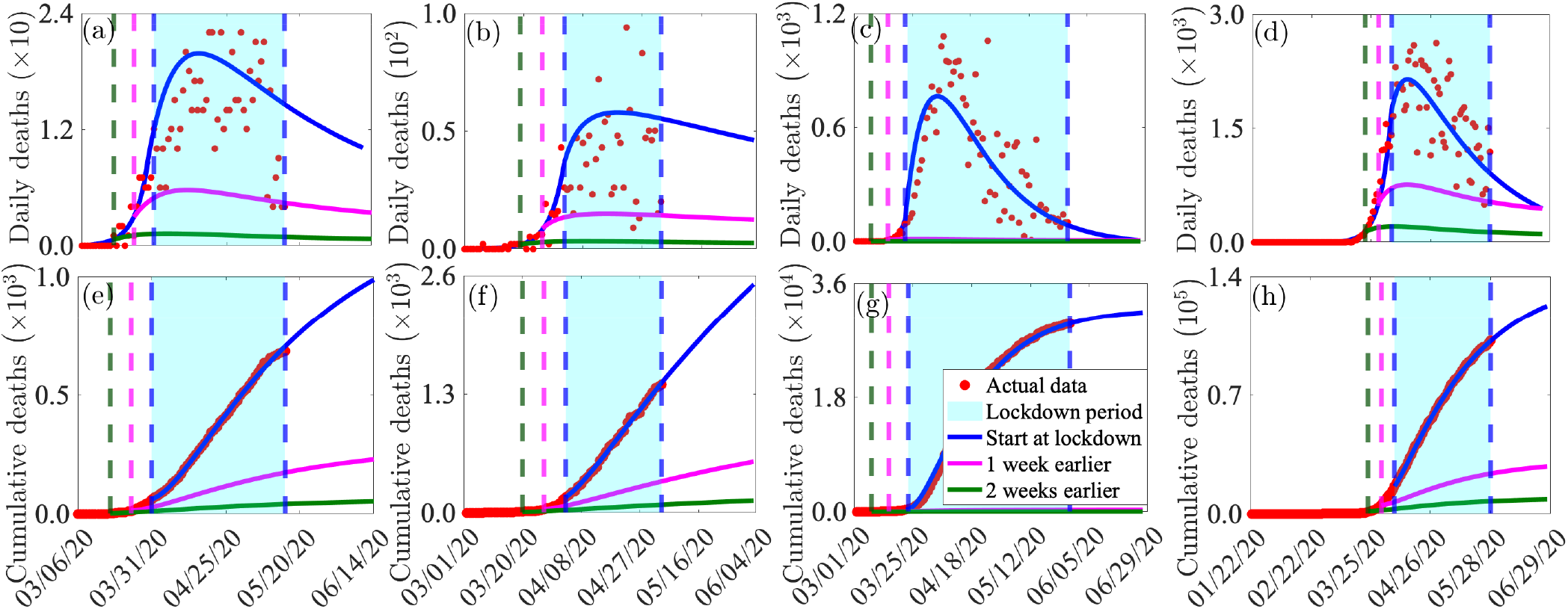
Impact of early lockdown: Simulations of the model (2.1), showing the daily and cumulative mortality, as a function of time, for various durations of the onset of lockdown measures for the states of Arizona, Florida, New York and the whole of US. The model is ran from the date of the index case, for each of the four jurisdictions, and extended one month beyond the first day of the relaxation (or partial lifting) of the lockdown measures. (a)-(d): daily deaths for the state of Arizona, Florida, New York and all of US, respectively. (e)-(h): cumulative deaths for the state of Arizona, Florida, New York and all of US, respectively. The predictions of the model, for the case when community lockdown measures were implemented one week earlier, two weeks earlier, or on the actual date the lockdown measures were implemented, are represented by magenta, green, and blue curves, respectively. Red dots represent the actual observed data, while blue dashed vertical lines depict the start and end of the actual lockdown period (shaded in cyan). Parameter used in the simulations are given in Tables A.3-A.7 in Appendix A, with various start dates for the implementation of community lockdown measures.

It is worth mentioning that, for the case of the state of New York for instance, the cumulative number of confirmed cases recorded during the week starting March 15, 2020 (i.e., a week before the lockdown measures were implemented in New York state, March 22, 2020) was 740. The cumulative number of deaths state-wide was 10 as of that week [4, 5]. These cumulative confirmed cases and deaths rose to about 15, 885 and 209, respectively, a week later [4, 5], demonstrating the exponential growth of the pandemic during that time. Our simulations show that the pandemic could have been suppressed in the state of New York if the lockdown measures were implemented during the week of March 14, 2020 (i.e., if they were implemented a week earlier). This result is consistent with the fact that many countries in Asia and South Pacific, such as China, Hong Kong, Japan, Taiwan, and New Zealand, successfully curtailed the burden of the COVID-19 pandemic by their early implementation of lockdown measures [51–55]. For instance, as of July 8, 2020, China (the first epicenter of COVID-19, and with a population of about 1.4 billion people) recorded only 83, 572 cumulative confirmed cases and 4, 634 cumulative mortality during the pandemic [4, 5]. Similarly, as of July 8, 2020, Japan (with a population of about 127*million*) recorded about 21, 000 cumulative confirmed cumulative cases and 980 cumulative mortality [4, 5].

Simulations were also carried out to assess the community-wide impact of increased face mask usage during the lockdown period in each of the four jurisdictions considered in this study. For these simulations, four levels of face mask compliance (in relation to the baseline value of the face mask compliance during lockdown, tabulated in Tables A.3-A.7 in Appendix A), namely 25%, 50%, 75% and 100% compliance, are considered. The results obtained, depicted in Figure 5, show a marked decrease in cumulative mortality with increasing compliance level of mask usage in each of the four jurisdictions. In particular, it can be seen that, if half of the residents of the state of Arizona consistently wear face mask in public during the lockdown period, up to 47% of the cumulative mortality recorded by the day of the partial lifting of the lockdown measures in the state (May 14, 2020) would have been averted (Figure 5 (a)). Similarly, up to 30%, 50% and 26% of the cumulative deaths recorded by the day of the lifting of the lockdown in the states of Florida (Figure 5 (b)), New York (Figure 5 (c)) and in the entire US (Figure 5 (d)) would have been averted if half their residents wear face masks consistently during their respective lockdown periods.

**Figure 5:**
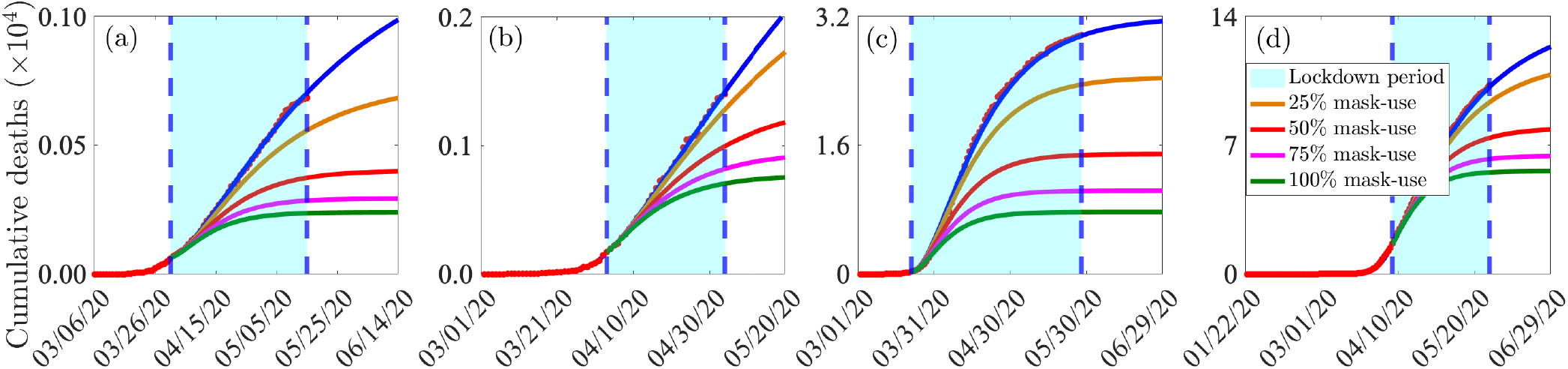
Impact of face masks compliance during lockdown: Simulations of the model (2.1), showing cumulative mortality, as a function of time, for the US state of Arizona, Florida, New York and the entire US. The model is simulated from the date of the index case for each of the four jurisdictions, for different levels of face masks compliance (*c*_*m*_). (a)-(d): cumulative deaths for the states of Arizona, Florida, New York and all of US, respectively. The blue curve represents the case for the baseline value of the face masks compliance, and the red dots represent the actual data. The blue dashed vertical lines represent the onset and termination dates for the lockdown measures (this region is shaded in cyan color). Parameter values used in the simulations are as given in Tables A.3-A.7 in Appendix A, with various levels of face masks compliance (*c*_*m*_).

A dramatic increase in the cumulative mortality averted in the four jurisdictions is recorded if 75% of the residents of the four jurisdictions wear face masks during the lockdown period. Specifically, while up to 60% and 42% of mortality recorded by the day of lifting of lockdown measures in Arizona and Florida would be averted, the state of New York and the entire US would prevent up to 65% and 37% of the deaths they recorded by the day of the lifting (if 75% of their respective residents wear face masks during the lockdown). If everyone in the four jurisdictions wear face mask during the lockdown, the percentage reduction of the cumulative mortality on the day of lifting of lockdown in the states of Arizona, Florida, New York and the whole of US further increase to 66%, 50%, 74% and 44%, respectively (this corresponds to 468, 714, 21, 928, and 44, 380 deaths being avoided in the respective jurisdictions, under this 100% face mask compliance scenario). The actual cumulative mortality recorded in the four jurisdictions by their respective day of the partial lifting of the lockdown (based on using the baseline values of the face mask compliance, given in Tables A.3-A.7 in Appendix A), together with the cumulative mortality that would have been recorded on the day of the lifting for various face mask compliance levels, are tabulated in Table 4.1.

**Table 4.1:**
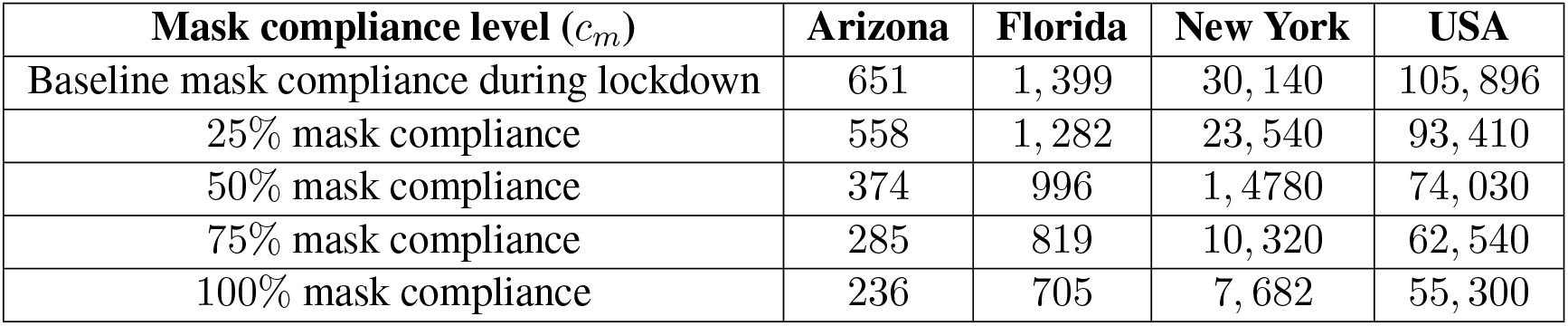
Actual cumulative mortality on the day lockdown was lifted (Row 2), and cumulative mortality as a function of mask use compliance that would have been recorded on the day lockdown was lifted? for the states of Arizona, Florida, New York, and entire US. Baseline mask compliance corresponds to the estimated mask-use compliance during the lockdown period in Tables A.3-A.7 in Appendix A.

| Mask compliance level ($c_m$) | Arizona | Florida | New York | USA |
| --- | --- | --- | --- | --- |
| Baseline mask compliance during lockdown | 651 | 1,399 | 30,140 | 105,896 |
| 25% mask compliance | 558 | 1,282 | 23,540 | 93,410 |
| 50% mask compliance | 374 | 996 | 1,4780 | 74,030 |
| 75% mask compliance | 285 | 819 | 10,320 | 62,540 |
| 100% mask compliance | 236 | 705 | 7,682 | 55,300 |

In summary, it can be concluded from Figures 4 and 5 that early implementation of lockdown (i.e., if they were implemented a week or two earlier than the day they were implemented), combined with increased level of face masks use during the lockdown periods, will result in a dramatic reduction in the burden of the pandemic in each of the four jurisdictions considered in this study. In fact, these figures show that it is quite possible that the pandemic might not have even (significantly) taken off in any of the four jurisdictions if the lockdown measures were implemented two weeks earlier, and if most of the residents of the four jurisdictions wore face masks consistently in public.

### 4.2 Impact of Varying Levels of Lifting of Community Lockdown Measures

The model (2.1) is now simulated, using the parameter values in Tables A.3-A.7 in Appendix A, to assess the community-wide impact of varying levels of lifting of the community lockdown measures implemented in the states of Arizona, Florida, New York and the entire US. Various states and jurisdictions within the US (with the exception of the states of New York, New Jersey, New Hampshire, District of Columbia, Illinois, and Delaware) implemented various levels of lifting of the lockdown measures (by May 20, 2020) [31], in an effort to re-open socio-economic activities. We consider three levels of community lockdown lifting, namely mild, moderate and high, as described below (adapted from [56]):

#### Mild re-opening

The main components of mild lifting or re-opening of community lockdown include lifting of stay-at-home (lockdown) measures (except for vulnerable individuals, who should shelter-at-home) and maintaining six-feet social-distancing in public places (e.g., parks, outdoor recreation areas, shopping areas, etc). For this re-opening level, no socializing in groups with more than 10 people and non-essential travels are allowed. Workers work from home (telework) and nonessential businesses (such as construction sites, manufacturers, nonessential retail stores for delivery, curbside and in-store pickup, wholesalers, outdoor business, such as drive-in movies, landscaping and gardening) are allowed to reopen. Furthermore, restaurants and bars are allowed to offer take-out, while schools and organized youth activities (e.g., daycare, camp) remain closed. Visitation to senior care facilities and hospitals are prohibited.

#### Moderate re-opening

Under this moderate level of community re-opening, indoor businesses (such as indoor dining at restaurants, hair salons and barber shops, but with waiting areas closed, offices, etc.,) are allowed to reopen (but with limits on capacity, strict cleaning requirements and mandatory social distancing). Furthermore, real estate firms, in-store retails (excluding Malls, but individual stores can provide curbside pickup), vehicle sales, leases and rentals, commercial building management, etc., are allowed to reopen. Nonessential travels and gatherings of up to 25 people are allowed. The use of face coverings in public is encouraged.

#### High re-opening

For high level of lifting of community lockdown, gatherings of up to 50 people are allowed. Additionally, nonessential businesses (such as indoor dining at restaurants with up to 50% capacity and tables spaced at least six feet away from each other; seating in bar areas allowed, but only if six feet of distance can be maintained between parties) are allowed to re-open. Furthermore, personal care services, such as nail salons, massage parlors, spar services and tanning salons can reopen. Some “low-risk” youth sports are allowed. Similarly, indoor religious gatherings can operate at 33% of maximum capacity. Nonessential businesses, such as, low-risk outdoor arts and entertainment activities, including zoos, botanical gardens, historical sites, outdoor museums and parks, low-risk indoor arts and entertainment activities, including museums, aquariums and historical sites; film, movie and music production; higher education institutions; professional sports, without fans in the stands, are allowed. Places that often draw large crowds (e.g., movie theaters, gyms and fitness centers, shopping malls, etc.,) are allowed to reopen.

We model lifting of community lockdown based on increases in community contacts, as measured in terms of increases in the baseline values of the community contact rate parameters, *β*_*p*_, *β*_*a*_, *β*_*i*_ and *β*_*s*_ (tabulated in Tables A.4-A.7 (b)). In particular, we assume that mild lifting of lockdown measures corresponds to a 5% increase in the baseline values of the contact rate parameters. Similarly, we assume that moderate and high lifting correspond to a 10% and 20% increase in the baseline values of these parameters, respectively. While numerous US states adopted mild or moderate levels of reopening, none, to the authors’ knowledge, adopted the high reopening level. In other words, high reopening represents a worst-case scenario that is not, at the current moment, realistically plausible. We do not expect any US state to adopt the high reopening level (until community transmission of the pandemic is greatly curtailed and/or a safe and effective vaccine or antiviral is available.

It should further be stated that, for the simulations to be carried out in this section, all control-related parameters of the model (e.g., parameters related to the use of face masks in public (*c*_*m*_ and *ε*_*m*_ and case detection (*τ*_*d*_, *τ*_*s*_ and *τ*_*dmax*_ etc.) are kept at their baseline values. That is, for the simulations in this section, no additional improvements in the baseline values of the control-related parameters of the model will be allowed (i.e., all control measures are implemented at their baseline levels given in Tables A.4-A.7).

The simulation results obtained, depicted in Figure 6, show an increase in both the daily and cumulative COVID-19-induced mortality with increasing lifting levels of the community lockdown. This is expected, since increasing the level of lifting of the community lockdown measures implies increased community contacts, thereby resulting in increased number of COVID-19 infections, hospitalizations and deaths. For the worst-case scenario, where the lifting level is high (i.e., the baseline values of the community contact rate parameters are increased by 20%), the states of Arizona, Florida and the entire US will experience a devastating second wave of the pandemic that will peak in about five to ten months after the lockdown measures were lifted, while the state of New York will only experience a mild second wave (Figure 6, red curves). In particular, under this high lifting scenario (and with control measures maintained at their lockdown baseline values), the second wave for the states of Arizona and Florida will peak in mid and late October, 2020, respectively, (with about 1, 148 and 2, 733 COVID-19 related deaths in Arizona and Florida, respectively, on the day the pandemic peaks (Figures 6 (a)-(b), red curves)). Similarly, the second wave for the entire US will peak early in February 2021 (recording about 2, 788 deaths on the day of the second wave peak), and no pandemic peak will be experienced at the state of New York (Figures 6 (c)-(d), red curves).

**Figure 6:**
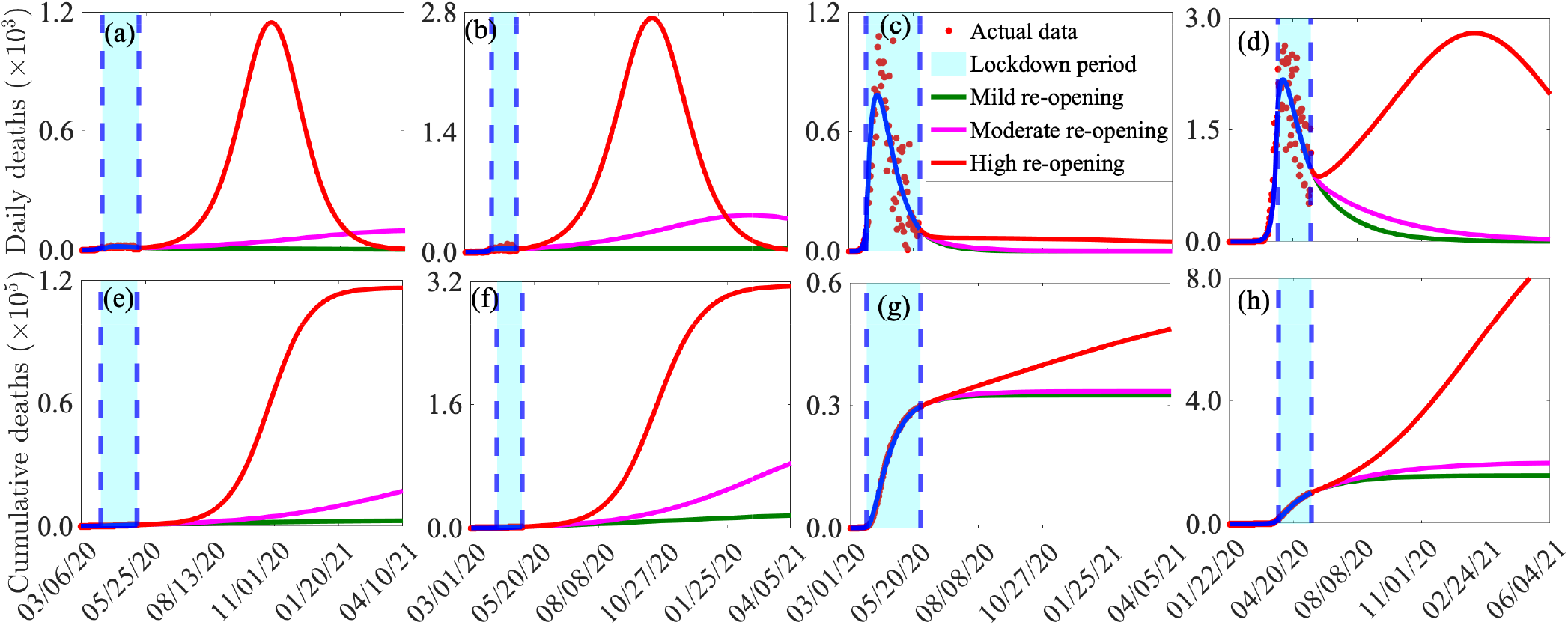
Impact of re-opening of community lockdown. Simulations of the model (2.1), showing daily and cumulative mortality, as a function of time, for various levels of re-opening of community lockdown measures in the states of Arizona, Florida, New York and the entire US. Mild, moderate, and high re-opening of community lockdown corresponds to a 5%, 10%, and 20% increase in the baseline value of the community contact rate parameters (*β*_*p*_, *β*_*a*_, *β*_*i*_, and *β*_*s*_), respectively. (a)-(d): Daily deaths for the state of Arizona, Florida, New York and all of US, respectively. (e)-(h): Cumulative deaths for the state of Arizona, Florida, New York and all of US, respectively. Other parameter values used in the simulations are as given in Tables A.3-A.7 in Appendix A.

On the other hand, if the level of lifting of the community lockdown was moderate, our results show a marked decrease in the size of the pandemic peaks, as well as a shift in the timing of the peaks (Figures 6 (a)-(d), magenta curves). Specifically, for this scenario, the peak of the second wave for Arizona and Florida will occur in mid to late April and February, 2021, respectively, while the US state of New York and the entire US do not experience a second wave. Furthermore, the moderate lifting scenario will avert over 92% and 84% of the deaths that would have occurred at the peak of the pandemic in Arizona and Florida, respectively, if high level of lifting of the control measures is implemented (i.e., compare red and magenta curves in Figures 6 (a)-(d)).

If mild lifting of the community lockdown was implemented, our simulations show that, while the states of Arizona and New York and the whole of US will not experience a second wave, the state of Florida will experience a relatively mild second wave that will peak in late November, 2020 (Figures 6 (a)-(d), green curves). In this (mild lifting) scenario, Florida will record about 46 deaths at the peak of the second wave. This represents a 98% reduction of the mortality that would have been recorded at the peak of the second wave in Florida if high lifting of lockdown measures was implemented. Furthermore, for this mild lifting of lockdown scenario, the states of Arizona, Florida, New York, and the entire US will record about 2, 274, 12, 06032, 380, and 155, 900 cumulative deaths, respectively, by the end of December 2020. These cumulative death numbers represent increases of about 438, 4, 252, 30, and 5, 300, respectively, in the cumulative mortality that would have been recorded by the end of September 2020. Additionally, the projected cumulative mortality numbers by the end of the year represent increases of about 1, 623, 10, 661, 2, 240, and 50, 004, respectively, from the actual cumulative mortality numbers when each of the four jurisdiction started re-opening (Figures 6 (e)-(h)). Table 4.2 summarizes the cumulative COVID-19 related deaths that would have been averted in the states of Arizona, Florida, New York, and the entire US, by the end of September, October, November, and December of 2020, if the mild level of lifting of community lockdown was implemented in the four jurisdictions, in comparison to implementing the moderate level of lifting of community lockdown (showing greater reductions in the states of Arizona and Florida, in comparison to the mild or moderate reductions in the state of New York and the whole of US).

**Table 4.2:** Percentage reduction in cumulative COVID-19 mortality averted in the states of Arizona, Florida, New York and the entire US (by the end of September, October, November, and December of 2020) if mild levels of lifting of community lockdown was implemented, in comparison to the case where moderate level of lifting of the community lockdown was implemented. Mild and moderate lifting of community lockdown correspond, respectively, to a 5% and 10% increase in the baseline values of the community contact rate parameters (*β*_*p*_, *β*_*a*_, *β*_*i*_ and *β*_*s*_).

| Location | September 30, 2020 | October 31, 2020 | November 30, 2020 | December 31, 2020 |
| --- | --- | --- | --- | --- |
| Arizona | 48 % | 58 % | 66 % | 73 % |
| Florida | 52 % | 60 % | 67 % | 72 % |
| New York | 2.3 % | 2.5 % | 2.6 % | 2.7 % |
| US | 10 % | 13 % | 15 % | 17 % |

It is worth emphasizing that the size of the second wave depends on the level of lifting of the lockdown measures implemented and the location. For instance, while the state of Florida will always record a second wave, regardless of the level of the lifting of the community lockdown measures (mild, moderate, or high), the states of Arizona and New York, as well as the entire US, will not record a (significant) second wave if the level of lifting of the community lockdown was mild. In all four jurisdictions, the severity (or pubic health burden) of the second wave decreases with decreasing level of the lifting of the community lockdown measures. Further, moderate and mild lifting not only decreases the peak daily mortality, they shift the time of the second wave. It is also notable from Figure 6 that the second wave in Arizona and Florida is far more severe than the first wave, regardless of the level of lifting of the community lockdown implemented in the two states. It is worth emphasizing from Figure 6 that, if the control measures are kept at their baseline levels during the lockdown periods for the states of Arizona and Florida (i.e., if no additional control measures, such as increased face mask use compliance and shutting down of large gathering places, are implemented), the spikes observed in the states of Arizona and Florida, that started during the middle of June 2020, may perhaps be considered to signal the beginning of the projected second wave in the two states, under the high lifting scenario (which are expected to peak by mid to end of October 2020, respectively).

### 4.3 Effect of Mask Usage After High Lifting of Community Lockdown Measures

In this section, the model (2.1) is simulated, using the parameter values in Tables A.3-A.7 in Appendix A, to assess the community-wide impact of mask usage during the post-lockdown period in the states of Arizona, Florida, New York and the whole of US. For these simulations, we consider high level of lifting of the community lockdown measures in all four jurisdictions (i.e., we consider the scenario where the baseline values of the community contact rate parameters, *β*_*p*_, *β*_*a*_, *β*_*i*_ and *β*_*s*_, are increased by 20% in all four jurisdictions) and various values of face mask compliance (*c*_*m*_). Note that the baseline efficacy of the face mask to protect the wearer from acquiring infection is assumed to be 50% (i.e., *ε*_*m*_ = 0.5). The simulation results obtained are depicted in Figure 7, from which it follows that the cumulative number of cases, hospitalizations and COVID-induced mortality decrease with increasing face mask compliance. It was observed that if two in every five people (i.e., 40%) in each of the jurisdictions considered in this study wear face masks in public after the lockdown measures were lifted, only the state of Florida will experience a (very mild) second wave of the pandemic. In this scenario, the states of Arizona and New York, as well as the entire US, will not have a second wave of the pandemic. Furthermore, no second wave of the pandemic will be experienced in any of the four jurisdictions if half of the residents wear face masks after the lockdown (this is obtained from plotting the corresponding daily mortality; the plot is not included to save space).

**Figure 7:**
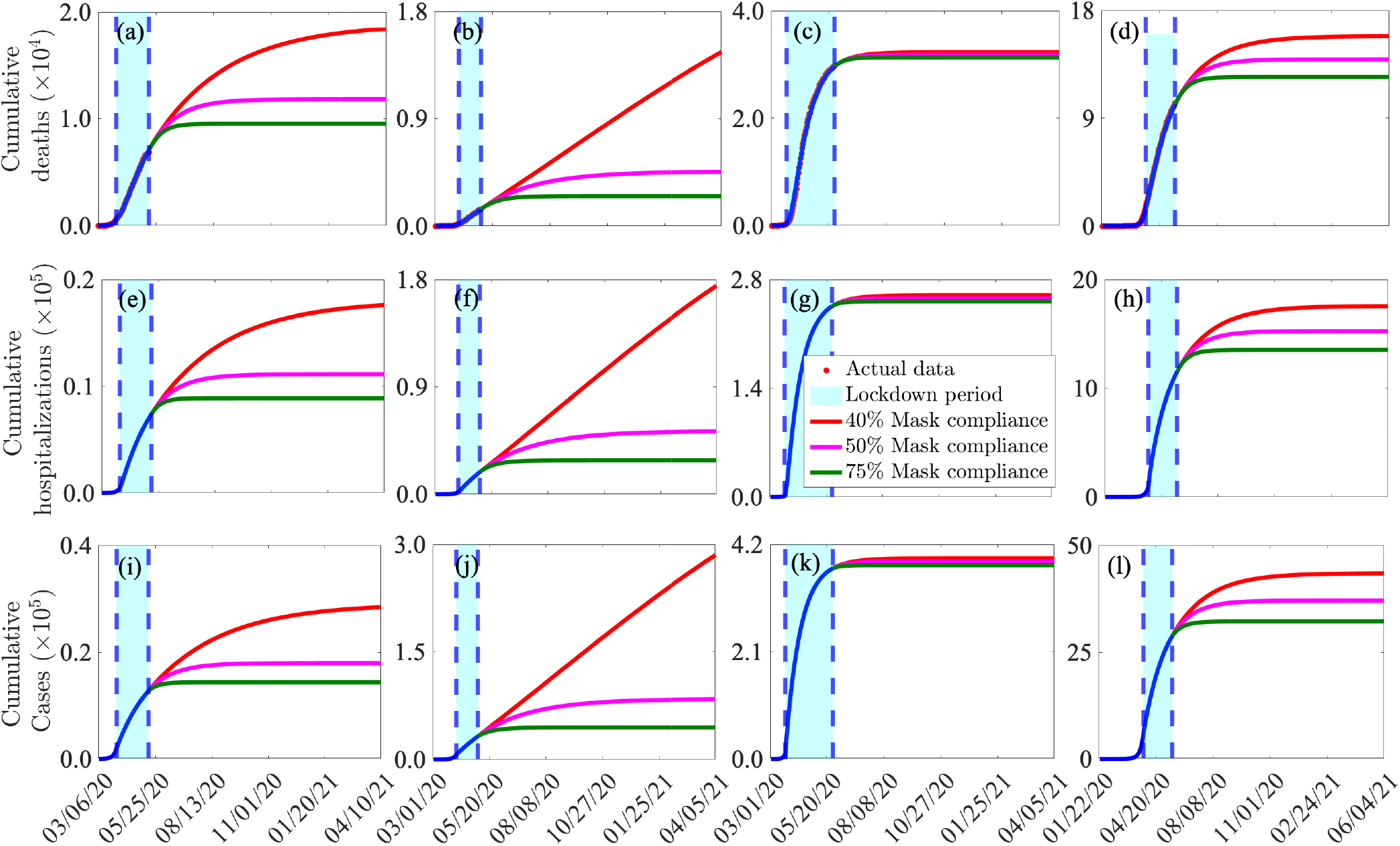
Impact of face mask usage on COVID-19 burden after high lifting of community lockdown. Simulations of the model (2.1), for high lifting of community lockdown in the states of Arizona, Florida, New York and the entire US, and various levels of face mask compliance (*c*_*m*_). (a)-(d): Cumulative mortality for the state of Arizona, Florida, New York and all of US, respectively. (e)-(h): Cumulative hospitalizations for the state of Arizona, Florida, New York and all of US, respectively. (i)-(l): Cumulative cases for the states of Arizona, Florida, New York and all of US, respectively. Parameter values used are as given in Tables A.3-A.7 in Appendix A, and various levels of face mask compliance (*c*_*m*_).

If half of Arizonans wear face masks after the lockdown period, under this high lifting scenario, Figure 7 (a) shows that the state will record a cumulative mortality of 1, 180 by the end of December 2020 (and this corresponds to a 98% reduction in the cumulative mortality that would have been recorded in the state if the face mask compliance was at the estimated value of 14% during the lockdown period, tabulated in Table A.4)). Since the face mask compliance level in Arizona immediately after the lockdown measures were lifted was (obviously) less than 50%, the actual cumulative mortality number for Arizona will be more than what we projected under the aforementioned 50% face masks use scenario. Nonetheless, the mandatory face mask use ordinances announced by many cities within the state in June 2020 [57] will significantly increase the face mask compliance, thereby helping to reduce the COVID-19 mortality in the state. Similarly, the states of Florida, New York, and the entire US will record 4, 465, 31, 770, and 138, 800 cumulative deaths, respectively (over the same time period), if half their populations wear face masks in public after the lockdown period. This represents a reduction of 96%, 26% and 67% in the cumulative mortality, respectively, that would have been recorded if the respective estimated face mask compliance during the lockdown period was used (Figures 7 (b)-(d)). These numbers decrease dramatically if 75% of the residents of the four jurisdictions wear face masks immediately after the lifting of the lockdown measures. For example, the states of Arizona, Florida, New York and the whole US will record cumulative mortality of 951, 2, 516, 31, 270 and 124, 200, respectively (over the same time period), if 75% of their respective residents wear face masks after the lockdown period (representing a 99%, 99%, 28% and 74% reduction in the respective cumulative mortality in comparison to when the estimated lockdown face mask compliance value is used). Higher reductions are observed when everyone wears face mask after the lockdown (*albeit* the reductions are far more pronounced in the states of Arizona and Florida, than in the state of New York and the entire US). Figure 7 shows similar results for the reductions in the cumulative hospitalizations (Figures 7 (e)-(h)) and the cumulative new COVID cases (Figures 7 (i)-(l)). Here, too, since the face mask compliance after the lifting of partial lockdown in Florida was less than 50%, the actual cumulative mortality in the state would be higher than projected under the 50% face mask use scenario. The compliance level is expected to increase following the announcement of mandatory face mask use ordinances in some cities and counties the state during June 2020 [58].

### 4.4 Effect of Mask Usage After High Lifting of Community Lockdown Measures

When high level of lifting is implemented (i.e., the associated community contact rate parameters (*β*_*p*_, *β*_*a*_, *β*_*i*_ and *β*_*s*_) are increased by over 20% from their baseline values), our simulations show that a second wave in all four jurisdictions would be inevitable, depending on the level of the increase of the high lifting scenario. We consider a hypothetical scenario where the level of lifting of the community lockdown measures is further increased, such as to a level that entails increasing the baseline values of the community contact rate parameters (*β*_*p*_, *β*_*a*_, *β*_*i*_ and *β*_*s*_) by 50%. This higher level may be akin to essentially returning to business as usual, where no significant restrictions are imposed (except, perhaps, no large gatherings such as major sporting, social and political campaign events). The simulation results obtained, for this hypothetical scenario in the four jurisdictions, are depicted in Figure 8. This figure shows a dramatic decrease in daily COVID-induced mortality with increasing face mask compliance. For this higher community lockdown lifting scenario, the states of Arizona and Florida will experience a major second wave peaking on October 19, 2020 (with 1, 294 deaths on this day) and October 3, 2020 (with 3, 599 deaths on this day), respectively, even if half their residents wear face masks after the lockdown has been lifted (Figures 8 (a)-(b), red curves). The state of New York and the entire US will have milder second waves peaking on March 24, 2021 (with 123 deaths at the peak) and January 13, 2021 (with 5, 896 deaths at the peak), respectively. Furthermore, no second wave will occur in any of the four jurisdictions if 75% of their respective residents wear face masks after the lockdown period (Figures 8 (a)-(d), purple curves). In fact, under this very high lifting scenario, all four jurisdictions will have very mild or no significant outbreaks of COVID-19 if the face masks compliance is at least 75%. In other words, high masks compliance after community lockdown lifting (even if the level of lifting was very high) greatly reduces the community transmission of COVID-19 in all four jurisdictions (resulting in very mild or no outbreaks in all four jurisdictions). If everyone in the jurisdictions wear face masks after the lockdown (i.e., *c*_*m*_ = 1), our simulations show that, for this very high level of lifting of community lockdown (with baseline community contact rates increased by 50%), the pandemic will be effectively curtailed (as measured in terms of major suppression of community transmission) in all four jurisdictions within two to three months after the community lockdown measures have been lifted.

**Figure 8:**
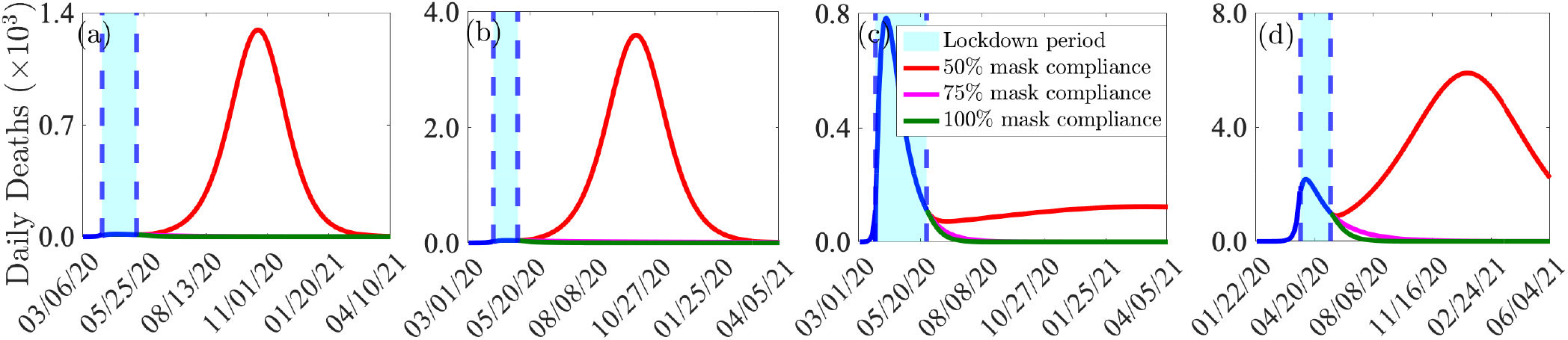
Impact of higher lifting of community lockdown measures. (as measured in terms of 50% increase in the baseline values of the community contact rate parameters, *β*_*e*_, *β*_*i*_, *β*_*a*_ and *β*_*s*_) and various compliance levels of face mask usage in the four jurisdictions. Simulations for the model (2.1) showing the daily mortality, as a function of time. (a)-(d): daily deaths for the state of Arizona, Florida, New York and all of US, respectively. Parameter values used are as given in Tables A.3-A.7 in Appendix A, with various levels of face masks compliance (*c*_*m*_).

### 4.5 Impact of Case Detection and High Lifting of Community Lockdown Measures

In this section, the model (2.1) is simulated to assess the impact of detection of exposed (*E*), pre-symptomatic (*E*_*p*_), and asymptomatically-infectious (*I*_*a*_) individuals, *via* the implementation of a COVID-19 diagnostic/surveillance testing strategy (as measured in terms of increase in the baseline values of the maximum detection rate parameter, *τ*_*dmax*_), and high level of lifting of community lockdown measures, on the burden of COVID-19 in the four jurisdictions considered in this study. The results obtained, depicted in Figure 9, show a dramatic decrease in the daily and cumulative mortality with increasing values of the maximum detection rate. This figure shows that, while Arizona, Florida, and the entire US will suffer a major second wave if the baseline value of the maximum detection rate (*τ*_*dmax*_) is used (as depicted in Figure 6), much milder second waves will be recorded in the states of Arizona and Florida if the baseline value of the maximum detection rate is increased by 10% (Figures 9 (a)-(d), red curves). It should be mentioned that the milder second waves will still be larger than the first waves recorded in the two jurisdictions. On the other hand, it can be seen from this figure that (for this scenario) the state of New York and the entire US will not suffer a second wave of the pandemic. If the baseline value of the maximum detection rate can be increased by 15%, our simulations show that none of the four jurisdictions will suffer a second wave. However, the decline in the daily deaths for the states of Arizona and Florida is slower that that for the state of New York and the entire US (Figures 9 (a)-(d), magenta curves). If the baseline value of the maximum detection rate is increased by 20% from its baseline value, our simulations show that none of the four jurisdictions will experience a second wave (Figures 9 (a)-(d), green curves).

**Figure 9:**
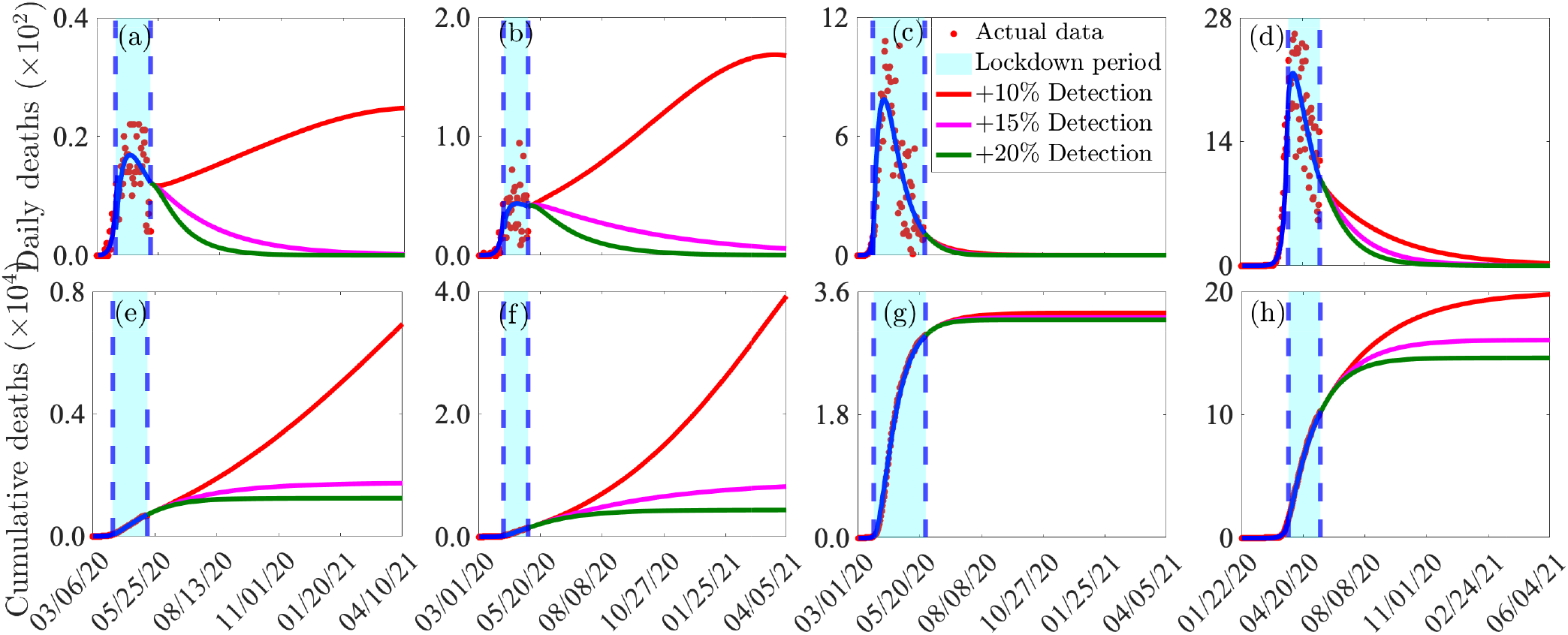
Impact of percentage increase in maximum detection rate of asymptomatic infected individuals, for the case where high level of lifting of community lockdown (i.e., 20% increase in the community contact rates) was implemented in each of the four jurisdictions. Simulations of the model (2.1), showing daily and cumulative mortality, as a function of time. (a)-(d): daily deaths for the state of Arizona, Florida, New York and all of US, respectively. (e)-(h): cumulative deaths for the state of Arizona, Florida, New York and all of US, respectively. Parameter values used are as given in Tables A.3-A.7 in Appendix A, with various percentage increase in the value of *τ*_*dmax*_.

Furthermore, a significant reduction in the cumulative mortality is achieved in all four jurisdictions, with increasing levels of the baseline maximum detection rate (Figures 9 (e)-(h)). For example, if the maximum detection rate is increased by 10% from its baseline value (noting that, for these simulations, high level of lifting of lockdown is used), our simulations for the state of Arizona show that up to 96% of cumulative mortality that would have been recorded by the end of December 2020, under the high lifting scenario with baseline value of maximum detection rate (depicted in Figure 6) will be averted in the state if *τ*_*dmax*_ is increased by 10% from its baseline value (i.e., compare Figures Figure 6 and Figure 9). Similarly, the states of Florida, New York and the entire US will avert 91%, 25%, and 60% of the respective cumulative deaths that would have been recorded by the end of December, 2020, under this scenario. Our simulations show even more dramatic decrease in the cumulative number of deaths if the baseline maximum detection rate is increased by 20% (Figure 9, green curves). It can be concluded from Figure 9 that widespread (random) diagnostic testing (i.e., the detection, tracing and rapid self-isolation of infected individuals with no symptoms of COVID-19) plays a major role in curtailing community transmission in each of the four jurisdictions. Consequently, widespread testing greatly contributes in suppressing community transmission of COVID-19. In particular, ramping up testing to a level that increases the baseline value of the maximum detection rate (*τ*_*dmax*_), by as low as 20%, could greatly diminish community transmission, to the extent that Arizona, Florida, New York state, and the entire US may not experience a second wave. In summary, widespread random testing contributes in detecting, tracing and isolating asymptomatic cases (hence, breaking their transmission chains) that would otherwise be spreading the virus in the community. More testing clearly does not mean more new cases. More testing means more detection (and rapid isolation) of asymptomatic cases, thereby reducing community transmission. It should be emphasized that face masks compliance is maintained at the baseline value depicted in Tables A.3-A.7.

Additional simulations were carried out to assess the combined impact of percentage increases in the maximum case detection rate (*τ*_*dmax*_) of asymptomatic infected individuals (i.e., infected individuals in the *E, E*_*p*_ and *I*_*a*_ classes) and increased mask use compliance (*c*_*m*_), from their baseline values, on the public health burden of the pandemic in the four jurisdictions. The simulations were carried out for the case where a high level of lifting of the community lockdown (i.e., 20% increase in the baseline values of the community contact rate parameters) was implemented in each of the four jurisdictions. The results obtained are depicted in Figure 10, from which it follows that the daily and cumulative mortality significantly decreases with increasing levels of the maximum case detection rate (from its baseline value). For instance, even a 10% increase in the baseline value of the maximum case detection rate could lead to a sizable reduction in the cumulative mortality in all four jurisdictions. Furthermore, a 20% increase in the baseline level of the maximum case detection rate can dramatically decrease the burden of the pandemic in each of the four jurisdictions even if combined with low level of face mask compliance, such as 25% face mask compliance (Figure 10, green curves). In fact, this combination (of 20% increase in baseline value of *τ*_*dmax*_ and 25% face mask compliance) can avert a second wave in each of the four jurisdictions. For this particular case (with 20% increase in baseline value of *τ*_*dmax*_ and 25% mask compliance), the states of Arizona, Florida, New York and the whole of US will record, respectively, about 1, 075, 3, 394, 31, 620, and 138, 000 cumulative deaths by the first week of December 2020. This corresponds to 99%, 99%, 28% and 71% decrease in the cumulative deaths if the maximum case detection rate (*τ*_*dmax*_) was combined only with baseline face mask compliance (i.e., if only baseline face mask compliance is used). Larger increases in the maximum case detection rate and increased face mask compliance will lead to even more significant decrease in the burden of the pandemic in all four jurisdictions.

**Figure 10:**
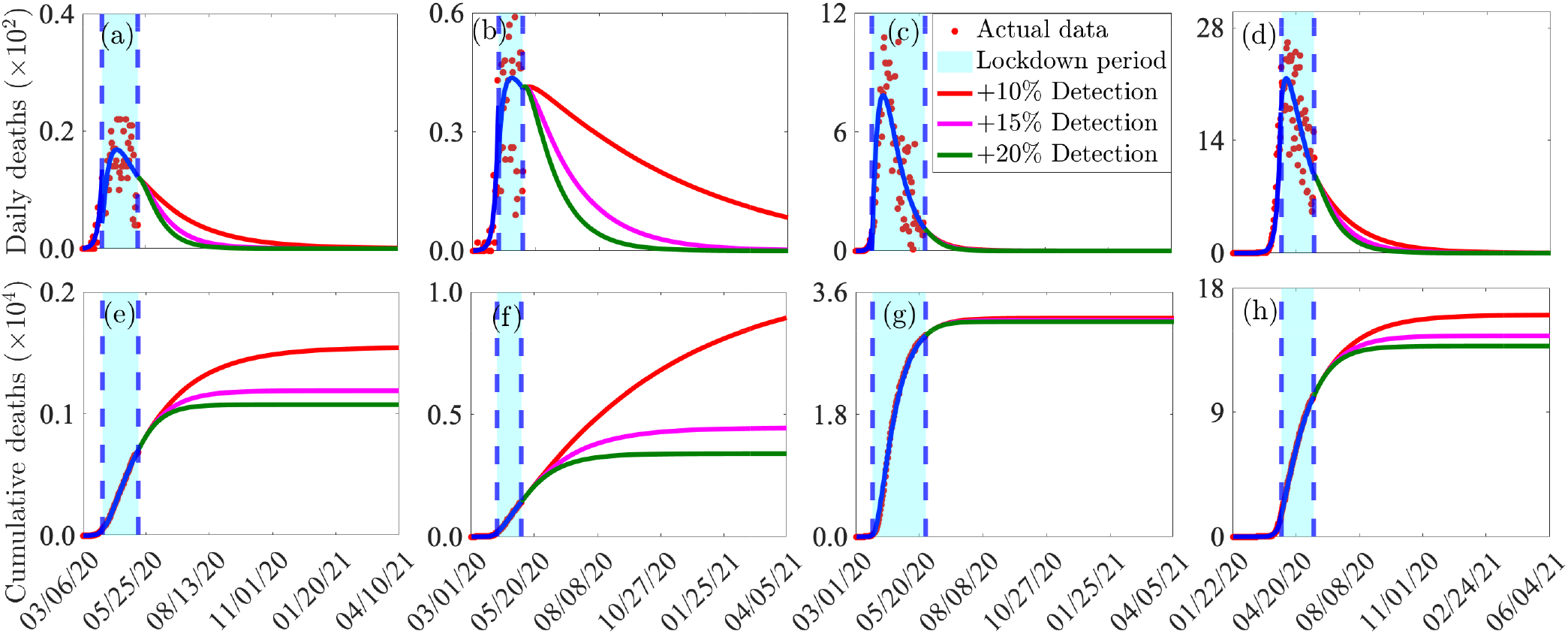
Impact of increase in maximum detection rate of asymptomatic infected individuals (*τ*_*dmax*_) and face masks use. Simulations of the model (2.1), showing daily and cumulative mortality, as a function of time, for high level of lockdown lifting in the states of Arizona, Florida, New York and the entire US. (a)-(d): daily deaths for the state of Arizona, Florida, New York and all of US, respectively. (e)-(h): cumulative deaths for the state of Arizona, Florida, New York and all of US, respectively. Parameter values used are as given in Tables A.3-A.7 in Appendix A, with various values of percentage increases in *τ*_*dmax*_ and face masks compliance fixed at 25% (i.e., *c*_*m*_ = 0.25).

## 5 Discussion and Conclusions

In this study, we developed a new Kermack-Mckendrick epidemic model (i.e., a disease transmission model with no demographic processes) for the transmission dynamics and control of COVID-19 in the states of Arizona, Florida, New York, and the entire US. Some of the notable features of the compartmental model, which takes the form of a deterministic system of nonlinear differential equations, include accounting for the dynamics of pre-symptomatic and asymptomatically-infectious individuals (who contribute to disease transmission) and allowing for the assessment of the community-wide impact of some non-pharmaceutical interventions, particularly the use of face masks in public. The model, which was parameterized using cumulative mortality data from the aforementioned four jurisdictions, was used to address the important question of whether or not the widespread use of face masks could halt the post-lockdown resurgence of COVID-19 in the US, without having to undergo another cycle of community lockdowns. Specifically, the model was used to assess the community-wide impact of early lockdown, various levels of lockdown lifting, case detection of asymptomatic individuals, and the use of face masks on the dynamics and control of COVID-19 in each of the four jurisdictions considered in this study.

Rigorous qualitative analysis of the model reveal that it has a continuum of disease-free equilibria, which is asymptotically-stable whenever a certain epidemiological threshold, known as the *control reproduction number* is less than unity. The epidemiological implication of this result, which represents a sufficient condition for the effective control of the disease, is that community transmission of COVID-19 can be significantly suppressed in the four jurisdictions if the control and mitigation interventions can bring, and maintain, the reproduction threshold to a value less than unity. A relation for the final size of the pandemic was also derived analytically.

Calibrating our model with the observed cumulative mortality data for the pre-lockdown and lockdown periods shows that pre-symptomatic (i.e., those in *E*_*p*_ class) and asymptomatic (i.e., those in *I*_*a*_ class) infectious individuals are the main drivers of the COVID-19 pandemic in each of the four jurisdictions (Table A.8). This can be intuitively justified based on the fact that pre-symptomatic and asymptomatic infectious individuals are typically not ill. Hence, they would continue to have the high community contacts they normally have, which can lead to their transmission of the disease to their susceptible contacts. The epidemiological implication of this result is that the rapid detection and isolation of infected individuals with no symptoms of the disease (as well as the rapid tracing and testing of their contacts) is critically-important to our ability to effectively control the pandemic (i.e., reducing community transmission, hospitalizations and deaths). Furthermore, our result (which is consistent with those reported in other studies, such as those in [21, 22]) suggests that even if all symptomatic individuals are detected and isolated from the actively-mixing population, the pandemic can still be sustained in the four jurisdictions by the pre-symptomatic and asymptomatically-infectious individuals in the jurisdictions. Again, this further reinforces the urgent need to detect and isolate infectious individuals who show no clinical symptoms of the disease.

We explored the sensitivity of the reproduction number with respect to face mask compliance in the four jurisdictions. In particular, we showed that community transmission of COVID-19 can be significantly reduced, using a face mask with efficacy of 50%, if at least 53% of Arizonans wear a face mask (from the beginning of the index case in Arizona). Similar face mask compliance for the states of Florida (59%), New York (38%) and the entire US (46%) were also obtained. In other words, our study shows that COVID-19 could have been effectively controlled if a public face mask strategy (particularly using surgical/medical masks, with protective efficacy of at least 50%) is implemented in each of the four jurisdictions, and the aforementioned minimum compliance attained (and maintained) from the beginning of the pandemic. This result is consistent with what actually happened in other parts of the world (particularly in some Asian countries, such as China, Japan, Singapore and South Korea), where the universal use of face masks greatly curtailed community transmission of COVID-19 (and brought the pandemic under very effective control, essentially at scales that can be, for all intents and purposes, considered as pandemic elimination).

Community lockdown was one of the major non-pharmaceutical intervention implemented in the US (and around the world), in an effort to curtail the community spread of COVID-19. Many states of the US were in community lockdown generally from mid March to the end of May, 2020 (we consider, for simulation purposes, that the entire US was (generally) in community lockdown between April 7, 2020 to May 28, 2020). In particular, the state of New York, once the global epicenter of COVID-19, was among the first of the US states to implement a community lockdown (on March 22, 2020) and among the last to begin to partially-reopen (on May 28, 2020). As a consequence, the state of New York is currently recording very low COVID-19 case numbers and one of the lowest mortality numbers in the US [59]. On the other hand, the states of Arizona and Florida that were among the last to lockdown and among the first to partially lift the lockdown measures, are now experiencing an alarming resurgence of COVID-19 (with record case and hospitalization numbers [4, 5, 60]. It should be emphasized that, although the states of Arizona and Florida succeeded in bending their epidemic curves during the lockdown period, they were not in community lockdown long enough to achieve the two weeks of continuous decline in confirmed cases stipulated in the re-opening guidelines by the White House Coronavirus Task Force [32]. Our study highlights the importance of early implementation of control measures (community lockdown in this case) and sustaining these measures until a time is reached when it is safe to (responsibly) begin to relax these measures (i.e., begin to partially lift the lockdown measures). Our simulations showed that if the lockdown measures were implemented a week earlier (than the actual days they were implemented in the four jurisdictions), all four jurisdictions would have recorded very significant reductions in their respective daily and cumulative mortality numbers by the day the lockdown measures were partially lifted. For example, our simulation results showed that about 77% of the cumulative mortality number for the US (as of the day lockdown measures were partially lifted) would have been averted. This result is consistent with the fact many countries in Asia and the South Pacific, notably China, Hong Kong, Japan, Taiwan and New Zealand, greatly succeeded in effectively halting the COVID-19 pandemic based on their very timely implementation of community lockdown control measures.

Our simulation results also highlight the importance of combining early implementation of the lockdown measures with increased face mask usage during the community lockdown. In particular, we showed that combining the early implementation of lockdown (i.e., implementing the lockdown measures 1-2 weeks earlier) with a public face mask strategy during the lockdown (and with reasonably high face mask compliance level) resulted in dramatic reductions in disease burden in each of the four jurisdictions. In fact, our simulations showed that the pandemic might not have even taken off significantly in any of the the four jurisdictions if the community lockdown measures were implemented two weeks earlier and most people in the jurisdictions wear face masks during the lockdown.

The lifting of community lockdown in many communities in the US was done in multiple phases, and at varying levels. For this reason, we used our model to assess the community-wide impact of the varying levels of lifting of community lockdown implemented in the four jurisdictions. We considered three levels of lifting of community lockdown, namely mild, moderate and high (based on the level of community interactions allowed. For example, based on whether or not restaurants, gyms, salons, malls, etc., are opened or closed). Mechanistically, the heterogeneity in the lifting levels is incorporated into our model by increasing the baseline values of the community contact rate parameters (*β*_*p*_, *β*_*a*_, *β*_*i*_ and *β*_*s*_). Our results showed an increase in daily and cumulative mortality in each of the four jurisdictions with increasing lifting levels of the community lockdown implemented in the respective jurisdictions. Furthermore, we showed that if a high level of lifting of the community lockdown is implemented (and all control measures are fixed at their lockdown period baseline levels), the states of Arizona and Florida will have a devastating second wave of the COVID-19 pandemic that will peak in about five months after their respective lifting of the community lockdown, while the state of New York and the entire US will record much milder second waves. However, if the level of lifting was moderate, our results showed a sizable decrease in the COVID-19 burden in all four jurisdictions. If mild lifting was implemented, only the state of Florida will experience a second wave of the pandemic. In other words, our study showed that the size (or severity) of the second wave depends on the level of lifting of the community lockdown. Furthermore, the severity (or size) of the second wave decreases with decreasing levels of the lockdown lifting. The second waves recorded in Arizona and Florida during moderate and high lifting of the community lockdown measures are drastically more severe than their respective first waves.

We also assessed the impact of universal use of face masks after the lockdown. In particular, we showed that for high lifting, the disease burden in each of the four jurisdictions decreases with increasing face mask compliance. If two in every five residents of each of the four jurisdictions (i.e., 40%) wear face masks after the lockdown, our study showed that only the state of Florida will have a second wave (which will be relatively mild). In fact, no second wave will be experienced in any of the four jurisdictions if half of their residents wear face masks. Higher reductions of pandemic burden will be recorded if face mask compliance is 75%. Thus, this study showed that, for mild moderate, or high level of lifting of community lockdown, face masks are extremely useful, and can greatly suppress COVID-19 (in addition to obviating the likelihood of a devastating second wave). On the other hand, if the level of lifting is unreasonably high, characterized by a 50% increase in the community contact rate parameters (this hypothetical scenario is essentially equivalent to returning to almost a “business as usual” scenario), then all four jurisdictions will experience a second wave (*albeit* the second wave for New York state and the entire US will be mild) if half the residents of the respective jurisdictions wear face masks after the lockdown. There will, however, be no second wave in any of the four jurisdictions if the post-lockdown face mask compliance is 75%. Thus, our study showed that high face masks compliance after lockdown will greatly curtail COVID-19 even if the lifting of the lockdown was high.

The community-wide impact of early detection and self-isolation of asymptomatic cases (as measured by increase in the maximum detection rate parameter, *τ*_*dmax*_) was also assessed in our study. Our simulation results showed a significant reduction in the daily and cumulative mortality with increasing values of the maximum detection rate, in all four jurisdictions. Although the states of Arizona and Florida, as well as the entire US, will suffer devastating second waves when their respective baseline value of the maximum detection rate parameter are used, ramping up testing (and contact tracing, followed by self-isolation of cases) to a level that increases the respective baseline value of the maximum detection rate by just 10% will result in a much milder second wave in Arizona and Florida, and no second wave in New York state and the whole of the US. Further increasing the baseline maximum detection rate to 20% of its baseline value will guarantee no second wave as well as trigger a substantial reduction in the number of deaths in any of the four jurisdictions. More dramatic reductions in disease burden are achieved if the testing strategy is combined with a universal face masks use strategy.

In summary, this study emphasizes the importance of early implementation of effective control strategies. In the absence of a safe and effective vaccine or antiviral, the control and mitigation of COVID-19 rely solely on the implementation of non-pharmaceutical interventions. While the lockdown measures implemented in the US have greatly curtailed community transmission of COVID-19 during the lockdown period, numerous states within the US federation rushed to pre-maturely re-open or lift the lockdown measures, triggering an alarming resurgence of COVID-19 in numerous states. We showed that the implementation of an effective public face mask strategy will greatly control community transmission in the states of Arizona, Florida, New York, and the entire US if the compliance level in each of the four jurisdictions is high enough (regardless of the level of lifting of lockdown implemented). We showed that such a face mask use strategy can avert the projected devastating second expected to hit these jurisdictions by the end of the year 2020. The effectiveness of face masks to curtail the burden of COVID-19 is enhanced if it is combined with an effective testing strategy that can increase the maximum detection rate (of asymptomatic infected individuals) in the community. This study showed that the prospect for the effective control of the post-lockdown resurging COVID-19 in the states of Arizona, Florida, New York and the entire US is very promising using a face mask strategy, if the mask strategy is universally adopted in the country (and compliance is at least moderate). Before a safe and effective vaccine and/or antiviral is developed and approved for use in humans, the use of face masks in public is, undoubtedly, the best and most effective way to curtail community transmission of the COVID-19 pandemic in the four jurisdictions we considered. Of course, our study is not advocating that face masks must be worn in public until the vaccine or antiviral is developed. They should be worn until community transmission is greatly suppressed. Once this is achieved, the use of face masks in public can be relaxed, as long as diagnostic testing and contact tracing can be ramped up to quickly identify and suppress any new post-mask outbreak in the community. Countries in Asia, particularly (and some in Europe) adopted this approach to great effect. Once their mask use strategy (and other NPIs, such as social-distancing) succeeded in suppressing community transmission, they relaxed the use of face masks in public but continue to be very vigilant (*vis a vis* the rapid identification and suppression of any future outbreaks). It should be mentioned that contact tracing is more feasible (and effective) when community transmission is already suppressed to a low (and manageable) level. It is logistically-easier (or feasible) to effectively trace a small number of cases, than a large number.

The US is currently (as of July 10, 2020) recording almost 72, 000 confirmed cases daily, and, as noted by Dr. Anthony Fauci during his testimony at a Senate Committee on June 30, 2020, the number is expected to rise to 100, 000 daily if the rising trend does not turn around. Tracing the contacts of these large number of cases will be an almost impossible undertaking. Our study emphasize the urgent need to significantly curtail community transmission (*via*, primarily, the universal use of face masks in public, complemented by social-distancing, avoiding large gatherings and the implementation of other non-pharmaceutical interventions). Once the community transmission is significantly decreased, the face masks use strategy can be relaxed, and tracing and testing can then become the main strategy to rapidly detect and contain any future COVID-19 outbreaks in the community. However, it is also expected that COVID-19 burden in the US will increase when schools reopen in the fall and the flu season kicks in.

Although some US schools are opting to fully reopen (i.e., adopt a full in-person learning schedule), while others are opting for a hybrid approach, which combines in-person and remote/online learning modules, each of these learning models is associated with a certain degree of risk of disease transmission in the school system (with the teachers, administrators, staff and anyone in the schools with underlying conditions etc. expected to bear the brunt of the disease burden). Thus, it really is imperative that any effort to reopen schools must be done safely (and based on what the data says). Jurisdictions that are experiencing high resurgence of the pandemic should think twice before fully reopening their schools. Communities that are experiencing such high resurgence may have to consider a second community lockdown, if they are unable to implement a face masks use strategy (with the required level of compliance) that can significantly suppress community transmission. For such communities, fully reopening their schools, coupled with the potential negative role the forth-coming flu season may have on the pandemic, can only exacerbates the dire situation they are in. Based on these facts, it is plausible to surmise that our projections for the pandemic burden in the four jurisdictions may be under-estimating the actual burden that will be recorded if the potential impact of school reopenings in the fall and the oncoming flu season are taken into account. Of course, these additional (anticipated) pandemic burden be effectively suppressed if effective control measures are implemented and sustained (at least throughout the fall of 2020). For instance, implementing the CDC guidelines on schools reopening [61] can certainly minimize the risks associated with the reopenings.

Finally, this study highlights the fact that widespread random testing contributes in detecting, tracing and isolating asymptomatic cases (hence, breaking their transmission chains) that would otherwise be spreading the virus in the community. More testing clearly does not mean more new cases. More testing means more detection (and rapid isolation) of asymptomatic cases, thereby reducing community transmission. Hence, more testing reduces number of new cases, hospitalizations and deaths. More dramatic reduction in COVID-19 burden (measured in terms of reduction in the number of new cases, hospitalizations and deaths) is achieved when the public face masks use strategy (with at least moderate compliance) is combined with a robust and effective random testing (and subsequent tracing and rapid isolation of cases) strategy in the community.

## Data Availability

Data used for the simulations is publicly available. The model was analyzed using MATLAB version R2020a.

## Acknowledgements

ABG acknowledges the support, in part, of the Simons Foundation (Awards #585022) and the National Science Foundation (Award #1917512). CNN acknowledges the support of the Simons Foundation (Award #627346).

## Appendices A Tables of variable descriptions, parameter descriptions, and parameter values

**Table A.1:** Description of the state variables of the model (2.1)

| State variable | Description |
| --- | --- |
| $S$ | Population of susceptible individuals |
| $E$ | Population of exposed (newly-infected but not infectious) individuals |
| $E_p$ | Population of pre-symptomatic (infectious) individuals |
| $I_a$ | Population of asymptotically-Infectious individuals |
| $I_m$ | Population of infectious individuals with mild or moderate clinical symptoms of COVID-19 |
| $I_s$ | Population of infectious individuals with severe clinical symptoms of COVID-19 |
| $I_i$ | Population of infectious individuals in self-isolation |
| $I_h$ | Population of hospitalized individuals |
| $I_c$ | Population of individuals in ICU |
| $R_u$ | Population of untested recovered individuals |
| $R_t$ | Population of recovered individuals who received serology (antibody) test |

**Table A.2:** Description of parameters of the model (2.1)

| Parameter | Description |
| --- | --- |
| $\beta_p(\beta_a)(\beta_m)(\beta_s)$ | Effective contact rate for individuals in the $E_p(I_a)(I_m)(I_s)$ class |
| $\varepsilon_m$ | Efficacy of face masks to prevent transmission and acquisition of infection ($0 < \varepsilon_m \leq 1$) |
| $c_m$ | Compliance in face mask usage in the community ($0 < c_m \leq 1$) |
| $\tau_d$ | Detection rate of asymptotically-infected individuals *via* random diagnostic/surveillance testing |
| $\tau_s$ | Detection rate of untested recovered individuals *via* serology (antibody) testing |
| $\tau_{dmax}(\tau_{smax})$ | Maximum diagnostic (serology) detection rate |
| $T_n$ | Average number of testing conducted *per* day |
| $\sigma_e$ | Progression rate from $E$ to $E_p$ class |
| $\sigma_p$ | Progression rate of pre-symptomatic exposed individuals to $I_a, I_m$ or $I_s$ class |
| $r$ | Proportion of individuals in the $E_p$ class who show no clinical symptoms of COVID-19 at the end of the incubation period (and move to the $I_a$ class) |
| $1 - r$ | Proportion of individuals in the $E_p$ class who show clinical symptoms of COVID-19 at the end of the incubation period (and move to the $I_m$ class, at a rate $g(1 - r)\sigma$, or to the $I_s$ class, at a rate $(1 - g)(1 - r)\sigma$) |
| $g$ | Proportion of individuals in the $E_p$ class who develop mild symptoms of COVID-19 at the end of the incubation period |
| $1 - g$ | Proportion of individuals in the $E_p$ class who develop severe symptoms of COVID-19 at the end of the incubation period |
| $f$ | Proportion of individuals in the $I_s$ class who are hospitalized |
| $1 - f$ | Proportion of individuals in the $I_s$ class who are self-isolated |
| $\rho_m$ | Self-isolation rate for individuals in the $I_m$ class |
| $(1 - f)\rho_s$ | Self-isolation rate for individuals in the $I_s$ class |
| $f\rho_s$ | Hospitalization rate for individuals in the $I_s$ class |
| $\xi_i$ | Hospitalization rate of self-isolated individuals |
| $\psi_h$ | ICU admission rate for hospitalized individuals |
| $\gamma_a(\gamma_s)(\gamma_h)(\gamma_i)(\gamma_c)$ | Recovery rate for individuals in the $I_a(I_s)(I_h)(I_i)(I_c)$ class |
| $\delta_s(\delta_h)(\delta_i)(\delta_c)$ | Disease-induced death rate for individuals in the $I_s(I_h)(I_i)(I_c)$ class |

**Table A.3:**
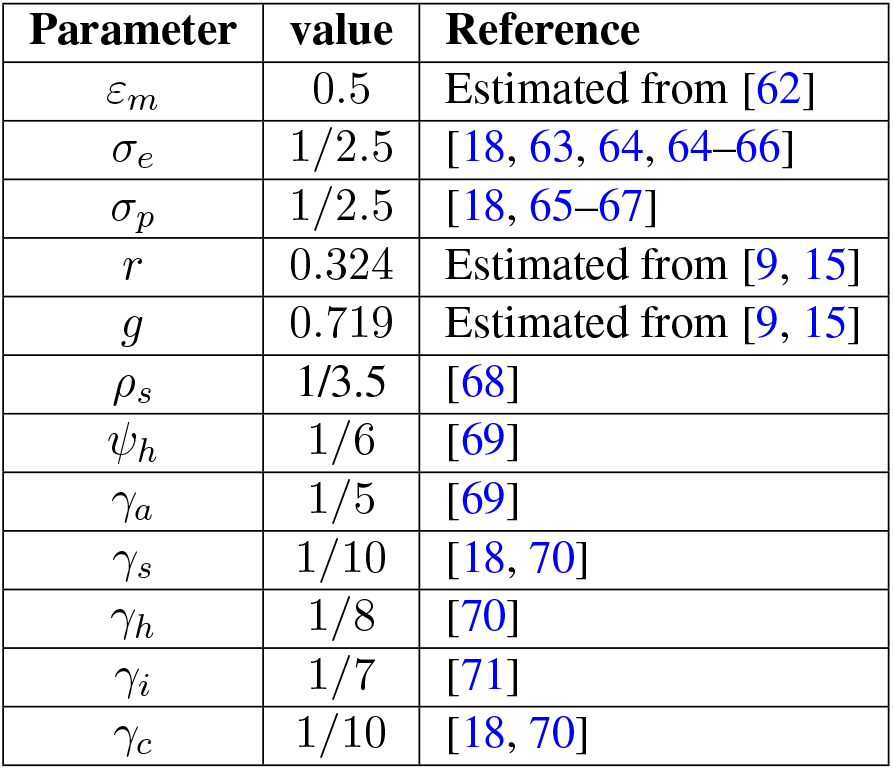
Baseline parameter values for the model (2.1) drawn from the literature.

**Table A.4:**
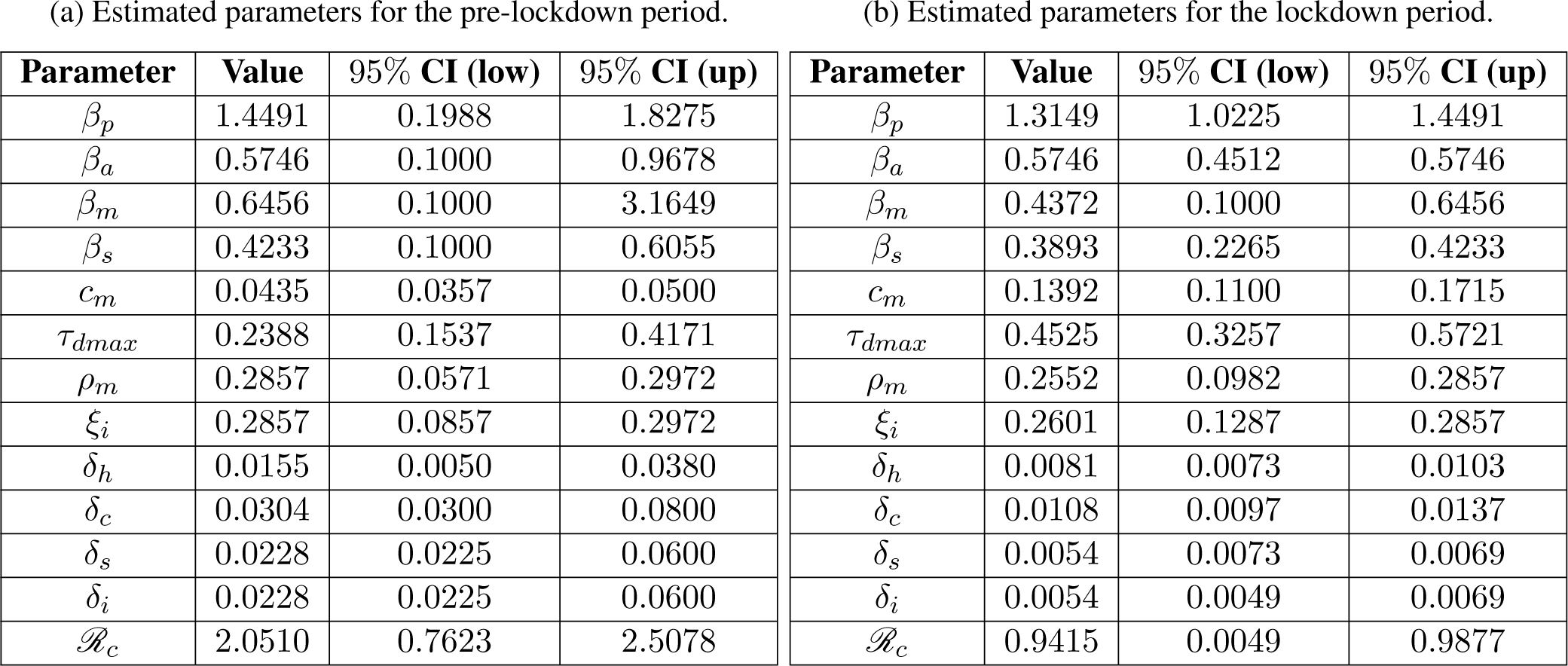
Calibrated parameter values for the model (2.1) using cumulative mortality data for the state of Arizona. (a) Pre-lockdown period (i.e., from March 6 to March 31, 2020). (b) Lockdown period (i.e., from March 31 to May 15, 2020). The lower 95% confidence interval bound is denoted by CI (low), while the upper 95% confidence interval bound is denoted by CI (up).

**Table A.5:**
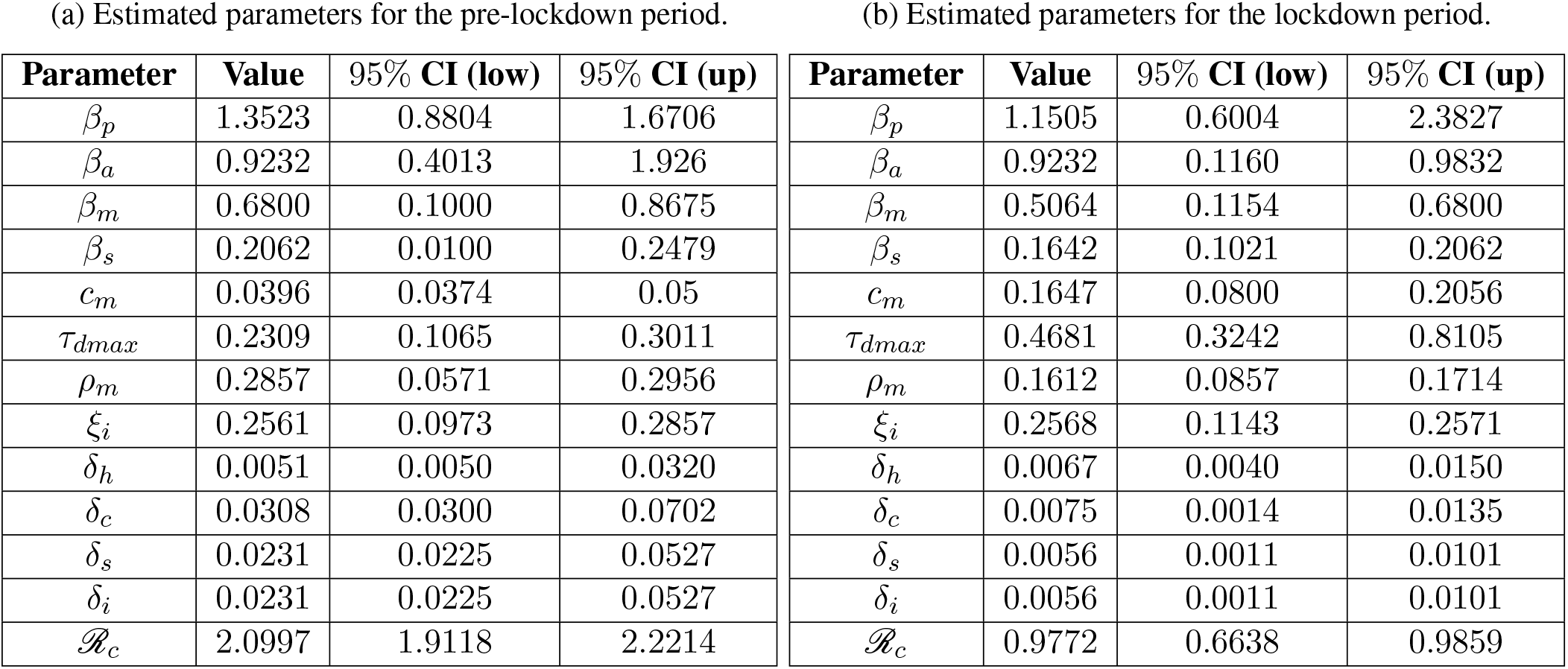
Calibrated parameter values for the model (2.1) using cumulative mortality data for the state of Florida. (a) Pre-lockdown period (i.e., from March 1 to April 3, 2020). (b) Lockdown period (i.e., from April 3 to May 4, 2020). The lower 95% confidence interval bound is denoted by CI (low), while the upper 95% confidence interval bound is denoted by CI (up).

| Parameter | Value | 95% CI (low) | 95% CI (up) | Parameter | Value | 95% CI (low) | 95% CI (up) |
| --- | --- | --- | --- | --- | --- | --- | --- |
| $\beta_p$ | 1.3523 | 0.8804 | 1.6706 | $\beta_p$ | 1.1505 | 0.6004 | 2.3827 |
| $\beta_a$ | 0.9232 | 0.4013 | 1.926 | $\beta_a$ | 0.9232 | 0.1160 | 0.9832 |
| $\beta_m$ | 0.6800 | 0.1000 | 0.8675 | $\beta_m$ | 0.5064 | 0.1154 | 0.6800 |
| $\beta_s$ | 0.2062 | 0.0100 | 0.2479 | $\beta_s$ | 0.1642 | 0.1021 | 0.2062 |
| $c_m$ | 0.0396 | 0.0374 | 0.05 | $c_m$ | 0.1647 | 0.0800 | 0.2056 |
| $\tau_{dmax}$ | 0.2309 | 0.1065 | 0.3011 | $\tau_{dmax}$ | 0.4681 | 0.3242 | 0.8105 |
| $\rho_m$ | 0.2857 | 0.0571 | 0.2956 | $\rho_m$ | 0.1612 | 0.0857 | 0.1714 |
| $\xi_i$ | 0.2561 | 0.0973 | 0.2857 | $\xi_i$ | 0.2568 | 0.1143 | 0.2571 |
| $\delta_h$ | 0.0051 | 0.0050 | 0.0320 | $\delta_h$ | 0.0067 | 0.0040 | 0.0150 |
| $\delta_c$ | 0.0308 | 0.0300 | 0.0702 | $\delta_c$ | 0.0075 | 0.0014 | 0.0135 |
| $\delta_s$ | 0.0231 | 0.0225 | 0.0527 | $\delta_s$ | 0.0056 | 0.0011 | 0.0101 |
| $\delta_i$ | 0.0231 | 0.0225 | 0.0527 | $\delta_i$ | 0.0056 | 0.0011 | 0.0101 |
| $\mathcal{R}_c$ | 2.0997 | 1.9118 | 2.2214 | $\mathcal{R}_c$ | 0.9772 | 0.6638 | 0.9859 |

**Table A.6:** Calibrated parameter values for the model (2.1) using cumulative mortality data for the state of New York. (a) Pre-lockdown period (i.e., from March 1 to March 22, 2020). (b) Lockdown period ((i.e., from March 22 to May 28, 2020)). The lower 95% confidence interval bound is denoted by CI (low), while the upper 95% confidence interval bound is denoted by CI (up).

| Parameter | Value | 95% CI (low) | 95% CI (up) | Parameter | Value | 95% CI (low) | 95% CI (up) |
| --- | --- | --- | --- | --- | --- | --- | --- |
| $\beta_p$ | 2.7450 | 1.3772 | 3.3895 | $\beta_p$ | 1.0363 | 0.4207 | 1.8901 |
| $\beta_a$ | 0.8989 | 0.1000 | 2.0134 | $\beta_a$ | 0.8989 | 0.2228 | 0.9889 |
| $\beta_m$ | 0.1254 | 0.1000 | 3.1228 | $\beta_m$ | 0.4046 | 0.1735 | 0.5978 |
| $\beta_s$ | 0.4246 | 0.1000 | 0.8043 | $\beta_s$ | 0.5943 | 0.1000 | 0.7425 |
| $c_m$ | 0.0190 | 0.0000 | 0.0500 | $c_m$ | 0.1500 | 0.1222 | 0.2162 |
| $\tau_{dmax}$ | 0.3010 | 0.3002 | 0.4940 | $\tau_{dmax}$ | 0.4485 | 0.3010 | 0.6142 |
| $\rho_m$ | 0.2857 | 0.0571 | 0.2896 | $\rho_m$ | 0.2466 | 0.1716 | 0.2857 |
| $\xi_i$ | 0.2822 | 0.0857 | 0.2857 | $\xi_i$ | 0.2426 | 0.1714 | 0.2571 |
| $\delta_h$ | 0.0371 | 0.0050 | 0.0400 | $\delta_h$ | 0.0097 | 0.0067 | 0.0221 |
| $\delta_c$ | 0.0434 | 0.0300 | 0.0800 | $\delta_c$ | 0.0129 | 0.0089 | 0.0221 |
| $\delta_s$ | 0.0326 | 0.0225 | 0.0600 | $\delta_s$ | 0.0065 | 0.0044 | 0.0111 |
| $\delta_i$ | 0.0326 | 0.0225 | 0.0600 | $\delta_i$ | 0.0065 | 0.0044 | 0.0111 |
| $\mathcal{R}_c$ | 2.5321 | 1.5460 | 3.2335 | $\mathcal{R}_c$ | 0.8480 | 0.5307 | 0.8955 |

**Table A.7:** Calibrated parameter values for the model (2.1) using cumulative mortality data for the entire US. (a) Pre-lockdown period (i.e., from January 22 to April 7, 2020). (b) Lockdown period (i.e., from April 7 to May 28, 2020). The lower 95% confidence interval bound is denoted by CI (low), while the upper 95% confidence interval bound is denoted by CI (up).

| Parameter | Value | 95% CI (low) | 95% CI (up) | Parameter | Value | 95% CI (low) | 95% CI (up) |
| --- | --- | --- | --- | --- | --- | --- | --- |
| $\beta_p$ | 0.7290 | 0.6677 | 0.7751 | $\beta_p$ | 0.4075 | 0.3327 | 0.5454 |
| $\beta_a$ | 0.8969 | 0.7745 | 0.9764 | $\beta_a$ | 0.6036 | 0.1008 | 0.8614 |
| $\beta_m$ | 0.8086 | 0.7530 | 0.9224 | $\beta_m$ | 0.4592 | 0.2531 | 0.7336 |
| $\beta_s$ | 0.4010 | 0.1870 | 0.4642 | $\beta_s$ | 0.1344 | 0.1053 | 0.2635 |
| $c_m$ | 0.0278 | 0.0276 | 0.0294 | $c_m$ | 0.1835 | 0.1312 | 0.2300 |
| $\tau_{dmax}$ | 0.1797 | 0.1722 | 0.1960 | $\tau_{dmax}$ | 0.2294 | 0.1799 | 0.3812 |
| $\rho_m$ | 0.1801 | 0.1584 | 0.1853 | $\rho_m$ | 0.2571 | 0.1045 | 0.2684 |
| $\xi_i$ | 0.1765 | 0.1316 | 0.2417 | $\xi_i$ | 0.0795 | 0.0571 | 0.1557 |
| $\delta_h$ | 0.0073 | 0.0052 | 0.0386 | $\delta_h$ | 0.0048 | 0.0043 | 0.0050 |
| $\delta_c$ | 0.0304 | 0.0300 | 0.0800 | $\delta_c$ | 0.0064 | 0.0058 | 0.0067 |
| $\delta_s$ | 0.0228 | 0.0225 | 0.0600 | $\delta_s$ | 0.0032 | 0.0029 | 0.0034 |
| $\delta_i$ | 0.0228 | 0.0225 | 0.0600 | $\delta_i$ | 0.0032 | 0.0029 | 0.0034 |
| $\mathcal{R}_c$ | 2.3277 | 2.2854 | 2.3758 | $\mathcal{R}_c$ | 0.8833 | 0.6363 | 0.8929 |

**Table A.8:** Percentage of community transmission generated by pre-symptomatic 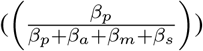 and asymptomatic infectious individuals 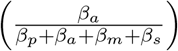 during the pre-lockdown and lockdown periods in the states of Arizona, Florida, New York, and the entire US.

| Period | Transmission source | Arizona | Florida | New York | US |
| --- | --- | --- | --- | --- | --- |
| Pre-lockdown | Pre-symptomatic individuals ($E_p$) | 47% | 43% | 65% | 26% |
| | Asymptomatic individuals ($I_a$) | 19% | 29% | 21% | 32% |
| Lockdown | Pre-symptomatic individuals ($E_p$) | 48% | 42% | 35% | 25% |
| | Asymptomatic individuals ($I_a$) | 21% | 34% | 31% | 38% |

